# A whole blood-based transcriptional risk score for nonobese type 2 diabetes predicts dynamic changes in glucose metabolic traits

**DOI:** 10.1101/2023.03.01.23286655

**Authors:** Yanan Hou, Huajie Dai, Na Chen, Zhiyun Zhao, Qi Wang, Tianzhichao Hou, Jie Zheng, Tiange Wang, Mian Li, Hong Lin, Shuangyuan Wang, Ruizhi Zheng, Jieli Lu, Yu Xu, Yuhong Chen, Ruixin Liu, Guang Ning, Weiqing Wang, Yufang Bi, Jiqiu Wang, Min Xu

**Affiliations:** Department of Endocrine and Metabolic Diseases, Shanghai Institute of Endocrine and Metabolic Diseases, Ruijin Hospital, Shanghai Jiao Tong University School of Medicine, Shanghai, China; Shanghai National Clinical Research Center for metabolic Diseases, Key Laboratory for Endocrine and Metabolic Diseases of the National Health Commission of the PR China, Shanghai Key Laboratory for Endocrine Tumor, State Key Laboratory of Medical Genomics, Ruijin Hospital, Shanghai Jiao Tong University School of Medicine, Shanghai, China

**Keywords:** nonobese type 2 diabetes, glucose metabolic traits, blood transcriptome, transcriptional risk score, risk prediction, transcriptome-wide Mendelian randomization

## Abstract

**Background:** The performance of peripheral blood transcriptional markers in evaluating the risk of type 2 diabetes (T2D) with normal weight is unknown. We developed a whole blood-based transcriptional risk score (wb-TRS) for nonobese T2D and assessed its contributions to disease risk and dynamic changes in glucose metabolism.

**Methods and findings:** We developed the wb-TRS in 1105 participants aged ≥40 years and in normal weight for up to 10 years from a well-defined community-based cohort with blood transcriptome data and validated it in an external dataset (253 overweight/obese participants from a dietary intervention trial with 3 repeated transcriptome data). Potential biology significance and causal inference were also explored. The wb-TRS included 144 transcripts. Compared to the lowest tertile, wb-TRS in tertile 3 associated with 8.68-folds (95% confidence interval [CI], 3.51-21.5), and each 1-unit increment associated with 2.57-folds (95% CI, 1.86-3.56) higher risk of nonobese T2D, after adjustments for traditional risk factors. Furthermore, baseline wb-TRS was significantly associated with dynamic changes in average, daytime, nighttime and 24h glucose and HbA1c, and area under the curve of glucose measured in the continuous glucose monitoring during 6-month of intervention. The wb-TRS improved the predicting performance for nonobese T2D in a model with fasting glucose, triglycerides and demographic and anthropometric parameters. Mitch analysis implicated oxidative phosphorylation, cholesterol metabolism and mTORC1 signaling involved in nonobese T2D pathogenesis. Transcriptome-wide Mendelian randomization supported causal effects of gene transcripts such as RAB1A and GCC1-PAX4 on nonobese T2D risk.

**Conclusions:** A whole blood based nonobese T2D associated TRS was validated to predict dynamic changes in glucose metabolism. These findings also suggested several genes and biological pathways that might involve in the pathogenesis of nonobese T2D.

## Author summary

**Why was this study done?**

- Blood transcriptome has been emerging as a potential novel approach to understand contribution of genes expression to biological functions and to identify key genes that confer to complex disease susceptibility, such as cardiovascular, psychiatry and chronic obstructive pulmonary disease.
- Rare studies have investigated the performance of the peripheral blood transcriptional markers in evaluating the risk and prediction of type 2 diabetes (T2D).

**What did the researchers do and find?**

- We developed a whole blood-based transcriptional risk score (wb-TRS) for T2D risk in a longitudinal community-based cohort.
- The wb-TRS was positively associated with risk of T2D and glucose metabolic traits. Furthermore, the baseline wb-TRS as well associated with dynamic changes of glucose metabolism measured in the continuous glucose monitoring in response to 6-month dietary intervention.
- The wb-TRS performed better and was complementary to fasting glucose, triglycerides and the demographic and anthropometric factors in predicting T2D.
- Transcriptome-wide Mendelian randomization and Mitch analysis and supported several gene transcripts (RAB1A and GCC1-PAX4) and biological pathways (oxidative phosphorylation, mTORC1 signaling, and cholesterol metabolism) that might involve in the pathogenesis of T2D.

**What do these findings mean?**

- Blood transcriptome highly related to and reflect risk of T2D and predict dynamic changes in glucose metabolism. It may lend insight into discerning potential causal genes and biological mechanisms in the pathogenesis of nonobese T2D.

## Introduction

Type 2 diabetes (T2D) has been a worldwide epidemic in recent decades leading high rates of early mortality and co-morbidity and possessing a considerable health and economic burden for the patients [1]. Early prediction and detection of individuals with glucose dysregulation and type 2 diabetes is extremely important for the prevention and management of these disorders. Previous reliable risk prediction models usually comprise the anthropometric measurements and traditional laboratory data. Recent studies have provided evidence of the prediction effect by including genetic factors and metabolic molecules in the models [2, 3]. However, as an essential biological and pathogenesis process, the performance and effect of gene transcription profile in evaluating T2D risk has not been well examined yet [4].

Tissue-specific gene expression (TSGE) is phenotypic manifestations integrating both genetic and environmental variations, which confers further insights to the intricate pathophysiology of diseases [5]. However, most tissues such as pancreas, liver, etc. cannot be readily obtained without invasive procedures in humans. As an alternative, blood transcriptome could better predict TSGE for ∼60% of the genes on average, with up to 81.7% in skeletal muscle, 71.5% in visceral adipose tissue, and 52.4% in pancreas [5]. Blood transcriptome has been emerging as a potential novel approach for complex diseases to understand contribution of genes expression to biological functions and to identify key genes that confer to complex disease susceptibility [6–9], such as cardiovascular, psychiatry and chronic obstructive pulmonary disease. However, rare studies have investigated the performance of the peripheral blood transcriptional markers in evaluating the risk and prediction of T2D.

Though obesity is one of its major risk factors, the T2D risk is still high in normal weight, especially in east Asians. One in three of diagnosed T2D had a body mass index (BMI) of less than 25 kg/m^2^, indicating the importance of metabolic health in the pathogenesis regardless of adiposity [10, 11]. Studies focused on refining T2D subtypes highlighting nonobese T2D, which characterized by a lower BMI, early onset of age, severe insulin deficiency, and serious complications [12–15]. The east Asians developed T2D risk at a relatively lower level of BMI [16]. The existing evidence indicated that specific genes, pathways, and biological mechanisms might underly the pathophysiology of nonobese T2D.

Thus, we developed a whole blood-based transcriptional risk score (wb-TRS) of nonobese T2D in 1105 participants aged 40 and above in an ongoing longitudinal Chinese cohort, in which the participants have maintained normal weight for up to 10 years. To validate its performance in a more general age and BMI range, we examined its predictive value on dynamic change in glucose metabolism in an independent external dataset (253 young overweight/obese participants from a dietary intervention trial with 3 repeated whole blood-based transcriptome data). Furthermore, we explored the potential genes and biological pathways that may conferred disease’s susceptibility and pathogenesis and performed transcriptome-wide Mendelian randomization (TWMR) to investigate additional evidence supporting a causal links between gene transcripts and T2D risk in a separate East Asian population and European population. The wb-TRS would provide an additional layer of biological interpretability for T2D.

## Methods

### Study population

The current analysis was based on one of the follow-up circles of our previous community-based cohort of in Shanghai, China [17–19]. Briefly, all the participants have taken part in a comprehensive examination that included a questionnaire and biochemical measurements at baseline in 2008-2009, the first (5-years) in 2013 and second (10-years) follow-up visits in 2018, respectively. The participants with known T2D before baseline examination were excluded.

According to the information at baseline and the 5-year follow-up examination, at the 10-year follow-up visit, we selected the participants whose BMI <25kg/m^2^ in past 10 years to take part in the examination sequentially and collected the whole blood samples for RNA-sequencing (RNA-seq). Finally, 1183 participants attended the follow-up visit finished the blood sampling for RNA-seq and 1105 with high-quality sequencing data were included in the present analysis. The flowchart of study participants was shown in **Fig S1**.

The study protocol was approved by the Institutional Review Board of Rui-Jin Hospital affiliated to Shanghai Jiao Tong University School of Medicine. All participants gave the written consents for the study.

### Data collection

A standard questionnaire was adopted to collect information on sociodemographic characteristics, disease and medical history, and lifestyles. Current smoking or drinking was defined as participants consumed cigarettes or alcohol regularly in the past 6 months. We used Global Physical Activity Questionnaire (GPAQ) to obtain frequency, duration, and intensity of physical activity. Physical activity level was evaluated as Metabolic Equivalent of Task (MET) minutes per week. Body height, weight and waist circumference were measured using a standard protocol. BMI was calculated as body weight (kg) divided by squared height (m^2^). Three seated systolic- and diastolic blood pressure (SBP and DBP) were measured (Omron HEM-752 Fuzzy; Omron, Dalian, China) at the non-dominant arm consecutively with 1 min interval after at least 5-minutes rest. The three readings were averaged for analysis.

### Biochemical measurements

Venous blood samples were obtained after an overnight fasting (>10 hours). All participants underwent a 75 g oral glucose tolerance test (OGTT). Fasting and 2h-OGTT glucose was measured using glucose oxidase method on an automated analyzer (ADVIA-1650 Chemistry System; Bayer); fasting and 2h-OGTT insulin using electrochemiluminescence assay on an automated analyzer (Roche Diagnostics, Basel, Switzerland); and hemoglobin A1c (HbA1c) using high pressure liquid chromatography on an automated analyzer (VARIANT II Hemoglobin Testing System, Bio-Rad Laboratories). Fasting triglycerides, total cholesterol, low-density and high-density lipoprotein cholesterol (LDL-C and HDL-C) were measured on the automated analyzer (ADVIA-1650 Chemistry System; Bayer).

### Definition of T2D

T2D was diagnosed as fasting glucose ≥7.0 mmol/l, 2h-OGTT glucose ≥11.1 mmol/l, HbA1c ≥ 6.5%, and/or taking antidiabetic therapy [20]. The homeostasis model assessment of insulin resistance (HOMA-IR) was calculated as fasting glucose (mmol/l) × fasting insulin (uIU/ml) / 22.5. The HOMA index for beta cell function (HOMA-β) was calculated as 20 × fasting insulin (uIU/ml) / (fasting glucose [mmol/l]-3.5).

### RNA-seq

The whole blood samples were collected by using the Paxgene RNA tubes according to the manufacture’s protocol, shipped and stored in −80℃ before RNA extracting. RNA was extracted by using the PAXgene Blood miRNA Kit Secondary BD Hemogard Closures (Catalog no 763134, Qiagen, Germany). Nanodrop One spectrometer (Thermo Fisher Scientific Inc., USA) and Qubit®3.0 Fluorometer (Life Technologies, CA, USA) were used to determine RNA purity and concentration. Isolated total RNA was analyzed on an Agilent Bioanalyzer 2100 (Agilent technologies, Santa Clara, CA, US) for RNA integrity number quality check. We excluded 78 samples with low RNA quality. 1ug RNA was used as input material for the RNA-seq preparations.

RNA-seq strand-specific libraries were constructed using the VAHTS Total RNA-seq (H/M/R) Library Prep Kit (Vazyme, China). Briefly, RNA was purified by magnetic beads after removal of rRNA. Then RNA was fragmented into small pieces using divalent cations for 8 minutes at 94℃. The cleaved RNA fragments were copied into first strand cDNA using reverse transcriptase and random primers. Second strand cDNA synthesis was subsequently performed using DNA Polymerase I and RNase H. These cDNA fragments then go through the end repair process, addition of a single ‘A’ base, and ligation of the adapters. The products were purified and enriched by PCR to create the final cDNA library. Purified libraries were quantified and validated by Qubit® 3.0 Fluorometer and Agilent 2100 bioanalyzer to confirm the insert size and calculate the mole concentration. Cluster was generated by cBot after the library diluted to 10 pM. Cluster were then sequenced in 150-bp paired-end mode on the Illumina NovaSeq 6000 platform (Illumina, USA) generating arounds 12 gigabases (Gb) reads per samples.

The base call and sequence quality across all generated FASTQ data was evaluated using FastQC (http://www.bioinformatics.babraham.ac.uk/projects/fastqc). We removed adapters, low quality reads, ribosome RNA reads, and reads with a sequence length less than 25 to obtain clean reads. Clean reads were mapped to human GRCh38.102 with Hisat2 (Hierarchical Indexing for Spliced Alignment of Transcripts, version 2.0.5) using default parameters. Overall, we obtained RNA-seq data with base mass (Q30) ≥90%, average mapped ratio was 94%. Count data were filtered to include transcripts with > 1count per million (CPM) in 99% of T2D cases or non-T2D controls.

### wb-TRS construction

The 1105 participants with blood-based transcriptome data were randomly assigned to training (75%) and testing (25%) dataset (R package caret) (**Fig 1**). To identify genes that could better distinguish the diseases status, we included the transcripts with gene expression level >1CPM in 99% of T2D cases or non-T2D controls and with a nominal difference P value <0.01 between these two groups in the wb-TRS features screening progress (n=757 gene transcripts). The wb-TRS construction was carried out in the training dataset and validated in testing and an external dataset. A multi-contrast gene set enrichment (Mitch) analysis [21] and Reactome pathway analysis [22] were performed to identify key genes and pathways.

**Fig 1.**
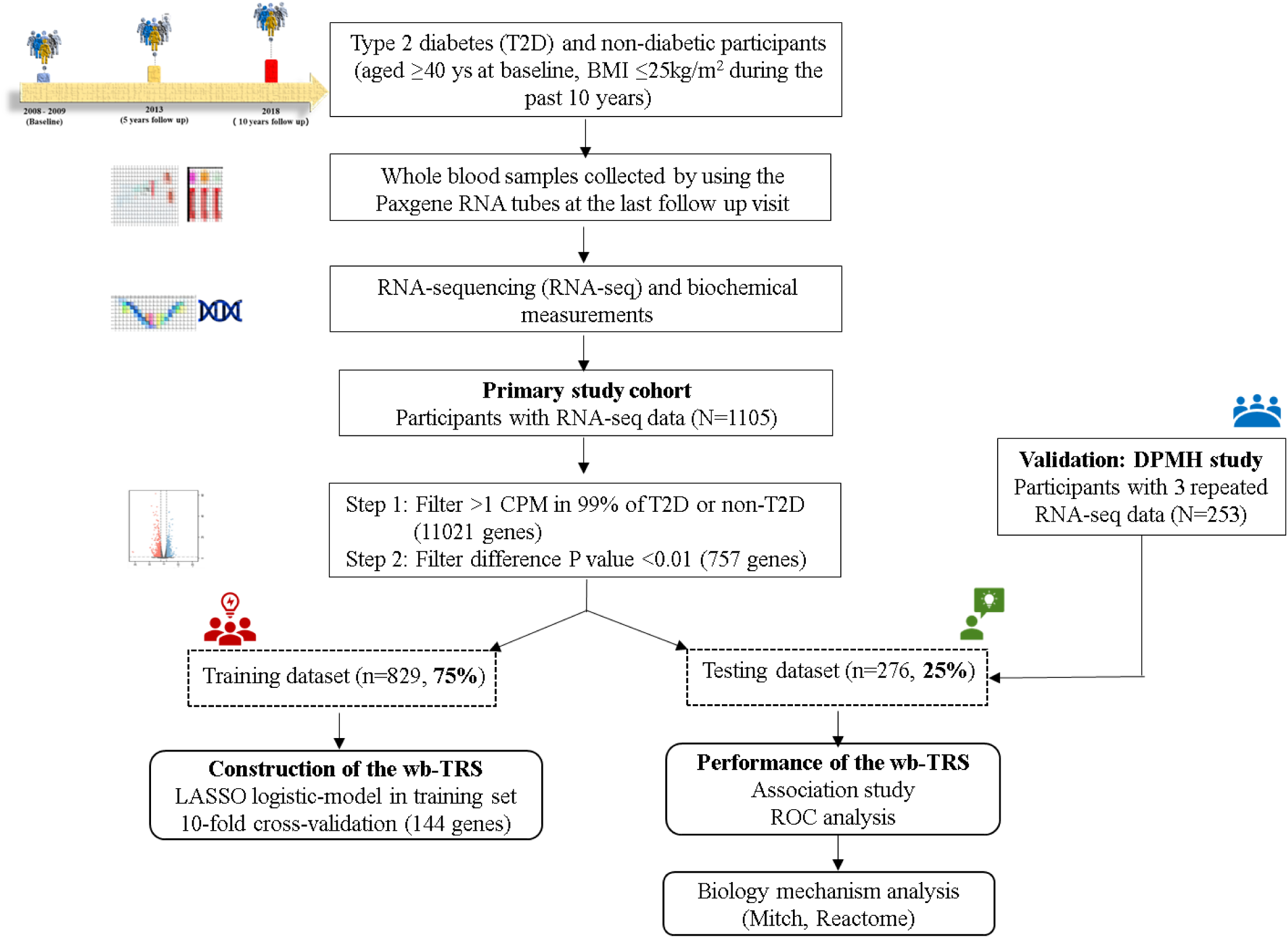
The schematic study design. At the final follow-up visit (10-year’s), the whole blood samples of the participants with BMI <25kg/m^2^ during the past 10 years were collected and underwent RNA sequencing (RNA-seq). The 1105 nonobese participants with RNA-seq data were randomly split into training (75%) and testing (25%) set. Transcripts with gene expression level >1 CPM in 99% of T2D cases or non-T2D controls and with a nominal difference P value <0.01 between these two groups in the wb-TRS features screening progress. In training dataset, we trained the LASSO logistic-model and adopted 10-fold cross-validation to conduct the transcriptional risk score (wb-TRS). In testing and external dataset, we tested the performance of wb-TRS. Finally, Mitch analysis and Reactome pathway analysis were performed to identify key genes and pathways. Abbreviations: T2D, type 2 diabetes; BMI, body mass index; CPM, counts per million; LASSO, least absolute shrinkage and selection operator; TRS, transcriptional risk score; Mitch analysis, multi-contrast gene set enrichment analysis.

In detail, we performed a penalized logistic regression framework using R package glmnet to develop the wb-TRS for nonobese T2D. We adopted the least absolute shrinkage and selection operator (LASSO) regression model to model training, which provides shrinks coefficients toward zero and automated feature selection. We adopted 10-fold cross-validation procedure to determine the lambda (0.006) that maximized the area under curve (AUC) for predicting T2D (**Fig S2**). The regression model with maximum AUC was used to calculate the wb-TRS.

### Statistical analysis

Multivariable logistic regression analyses were used to assess the risk of nonobese T2D in relation to wb-TRS. Odds ratio (OR) and the corresponding 95% confidence interval (CI) were calculated. The linear regression model was to assess the association of wb-TRS with the cardiometabolic traits. The restricted cubic spline (RCS) analysis was used to test any nonlinear associations between wb-TRS and glucose metabolism. The receiver operator characteristic curves (ROC) were used to evaluate and compare the predictive values of 6 models for nonobese T2D (R package pROC) including: 1. primary model (age, sex, BMI and family history of diabetes); 2. wb-TRS model; 3. fasting glucose and triglycerides model; 4. primary model adding fasting glucose and triglycerides (full); 5. primary model adding wb-TRS; 6. primary model adding fasting glucose and triglycerides (full), and wb-TRS.

### Independent validation

To validate the predictive value of wb-TRS for the dynamic changes in glucose metabolism, we performed analysis using data of 3 repeated blood-based transcriptome data and glucose metabolism in the Dietary Pattern and Metabolic Health (DPMH) study, a parallel-arm randomized controlled feeding trial conducted in 2019 (URL: https://www. clinicaltrials.gov; Unique identifier: NCT03856762) [23]. A total of 253 overweight/obese participants (aged 25 to 60 years, BMI ≥ 24.0 kg/m^2^, fasting glucose ≥ 5.6 mmol/l) with transcriptome data and glucose metabolic traits at baseline, the first (3-months) and second (6-months) follow-up visits were included in the validation analysis. The whole blood RNA was collected and underwent RNA-seq following the same procedure with the above cohort study. We adopted generalized estimating equations (GEE) models were to assess the association of three-repeated measurements of wb-TRS with three-repeated measurements of the glucose metabolism during the 6-months intervention. We further fixed the generalized linear models to assess the effects of baseline wb-TRS on dynamic changes of glucose metabolism at 3- and 6-months of intervention.

### The biological pathway and mechanism analysis

To gain biological insight behind the wb-TRS, we performed several bioinformatics analyses. We divided the participants in the initial cohort into 3 subgroups based on wb-TRS tertiles. We performed a multi-contrast gene set enrichment analysis by using R package mitch (v.1.4.0). The Mitch analysis uses a rank-MANOVA statistical approach to identify sets of genes that exhibit joint enrichment or divergent responses across multiple contrast. The input data was the log_2_(fold change) of all detected transcripts (n=54,458, including protein coding gene, lncRNA, and snRNA) for comparison between the bottom and mid tertile, and the top and mid tertile, respectively. The input genesets provided were 50 genesets: h.all.v7.5.1.symbols.gmt in Molecular Signatures Database (MSigDB). Enrichment score (ES) was obtained and ranged from -1 to 1. Meanwhile, we used the 144 gene transcripts included in the wb-TRS as input profiles to perform Reactome pathway analysis on reactome website (https://reactome.org/PathwayBrowser/#TOOL=AT).

### Transcriptome-wide Mendelian randomization

We performed a two-sample transcriptome-wide Mendelian randomization (TWMR) to identify causal associations of gene transcripts included in wb-TRS with T2D risk using the TwoSampleMR. The summary statistics of genome-wide association study of T2D was from the Asian Genetic Epidemiology Network (AGEN) [16] and the DIAbetes Genetics Replication And Meta-analysis (DIAGRAM) [24], respectively. We selected the significant expression quantitative trait loci (eQTLs with P <5×10^-^8) obtained from eQTLGen Consortium [25] and with linkage disequilibrium (LD) based on r^2^ <0.01 as valid instruments variants (IVs) used in TWMR. When no shared eQTLs were available between exposures and outcomes, proxies with LD r^2^ >0.8 were substituted. TWMR causality tests were performed using the inverse variance weighted method by meta-analyzing the effect of each IV from Wald ratio. Heterogeneity and pleiotropy of the results were assessed using Cochran’s Q statistic or MR-Egger intercept, respectively.

All analyses were performed using SAS (version 9.3; SAS Institute, Cary, NC, USA) and R software (version 3.5.1; R Core Team). A two-sided P value < 0.05 was considered statistically significant.

## Results

### Clinical characteristics of study participants

In the 1105 participants, 305 were defined as nonobese T2D (27.6%), 737 (66.7%) were women, with an average age of 64.5 (standard deviation [SD], 7.0) years old. The balanced clinical characteristics between the training and testing dataset indicated a low risk of selection bias introduced by random sampling (**Table S1)**.

### Construction and distribution of wb-TRS

Finally, 144 important features (gene transcripts) were included in the wb-TRS, which weights were displayed in **Table S2**. The wb-TRS was estimated as gene expression level of transcript1 × weight1 + gene expression level of transcript2 × weight2 +…+ gene expression level of transcript144 × weight144.

The wb-TRS was positively associated with age (β 0.98, SE 0.15, P<0.0001), and was slightly higher in men (−1.7, SD 1.5) than that in women (-2.1, SD 1.3). The mean wb-TRS was -0.5 (SD 0.9) in T2D and -2.5 (SD 1.1) in non-diabetes in training dataset, and -0.9 (SD 1.4) in T2D and -2.4 (SD 1.3) in non-diabetes in testing dataset. **Fig S3** displays the violin plot of the wb-TRS and gene transcripts most negatively (FUOM) and positively (RPF1) associated with T2D in training dataset.

### Association of wb-TRS with nonobese T2D

**Table 1** shows association of wb-TRS with nonobese T2D risk in testing dataset. Each 1-unit increment in wb-TRS (one score) was associated with a 2.49-folds higher risk of nonobese T2D (95%CI 1.84-3.36), after adjustments of age, sex, BMI and family history of diabetes. Additional adjustment for potential confounders including lifestyles, education levels, waist circumference, SBP and DBP, and lipids profiles did not change the results appreciably (OR 2.57, 95% CI 1.86-3.56). Specifically, each 1-unit increment in wb-TRS was associated with 3.40-folds higher risk of newly diagnosed nonobese T2D (95%CI 2.00-5.81), and 2.33-folds higher risk of diabetes with duration ≥ 5 years (95%CI 1.56-3.48). Compared with the bottom tertile, the middle or top tertile of wb-TRS was associated with 2.46- (95% CI 0.95-6.35) and 8.68-folds (95% CI 3.51-21.5) higher risk of nonobese T2D.

**Table 1.** Adjusted ORs (95% CI) of type 2 diabetes associated with whole blood-based transcriptional risk score (wb-TRS) in testing dataset. Data were odds ratio (OR), 95% confidence interval (CI) calculated from the multivariable logistic regression models. Model 1: adjusted for age, sex, body mass index, and family history of diabetes; Model 2: further adjusted for waist circumference, smoking and drinking status, education level, physical activity, systolic and diastolic blood pressure, total cholesterol, triglycerides, HDL-C, and LDL-C. Abbreviation: HDL-C, high-density lipoprotein cholesterol; LDL-C, low-density lipoprotein cholesterol.

|  | Tertile 1(-7.0 ~ -2.5) | Tertile 2 (-2.5 ~ -1.4) |  | Tertile 3 (-1.4 ~ -3.9) |  | P<sub>for trend</sub> | Each 1-unit |  |
| --- | --- | --- | --- | --- | --- | --- | --- | --- |
|  | / | OR (95% CI) | P value | OR (95% CI) | P value |  | OR (95% CI) | P value |
| Type 2 diabetes |  |  |  |  |  |  |  |  |
| Cases, n (%) | 8 (9.0) | 21 (23.3) |  | 47 (48.5) |  |  | 76 (27.5) |  |
| Model 1 | 1.00 (Ref.) | 2.57 (1.05-6.30) | 0.04 | 8.36 (3.56-19.7) | <.0001 | <.0001 | 2.49 (1.84-3.36) | <.0001 |
| Model 2 | 1.00 (Ref.) | 2.46 (0.95-6.35) | 0.06 | 8.68 (3.51-21.5) | <.0001 | <.0001 | 2.57 (1.86-3.56) | <.0001 |
| Newly diagnosed type 2 diabetes |  |  |  |  |  |  |  |  |
| Cases, n (%) | 2 (2.3) | 13 (14.4) |  | 19 (19.6) |  |  | 34 (12.3) |  |
| Model 1 | 1.00 (Ref.) | 6.41 (1.38-29.9) | 0.01 | 17.2 (3.72-79.8) | 0.0003 | <.0001 | 2.87 (1.84-4.48) | <.0001 |
| Model 2 | 1.00 (Ref.) | 6.77 (1.30-35.3) | 0.02 | 20.6 (3.94-108) | 0.0003 | 0.001 | 3.40 (2.00-5.81) | <.0001 |
| Type 2 diabetes with duration $\geq 5$ years | | | | | | | | |
| Cases, n (%) | 6 (6.7) | 7 (7.8) |  | 27 (27.8) |  |  | 40 (14.5) |  |
| Model 1 | 1.00 (Ref.) | 1.08 (0.33-3.58) | 0.90 | 5.74 (2.09-15.8) | 0.001 | 0.0002 | 2.32 (1.62-3.34) | <.0001 |
| Model 2 | 1.00 (Ref.) | 1.17 (0.33-4.24) | 0.81 | 6.12 (2.00-18.7) | 0.002 | 0.0001 | 2.33 (1.56-3.48) | <.0001 |

We then examined the associations of wb-TRS with nonobese T2D risk stratified by several traditional risk factors of T2D (**Fig S4**). A stronger association was identified in age <65 years and women (a marginal interaction between TRS and sex was observed with P for interaction =0.051).

### Association of wb-TRS with glucose metabolism

In both training and testing dataset, the wb-TRS was positively associated with fasting glucose, 2h-OGTT glucose, and HbA1c; but inversely associated with HOMA-β, in the fully adjusted model (**Table 2**). The RCS analysis showed similar results, except for that it demonstrated a J shaped relationships between wb-TRS and HbA1c and logHOMA_β (all P for non-linear ≤0.001**) (Fig 2)**.

**Fig 2.**
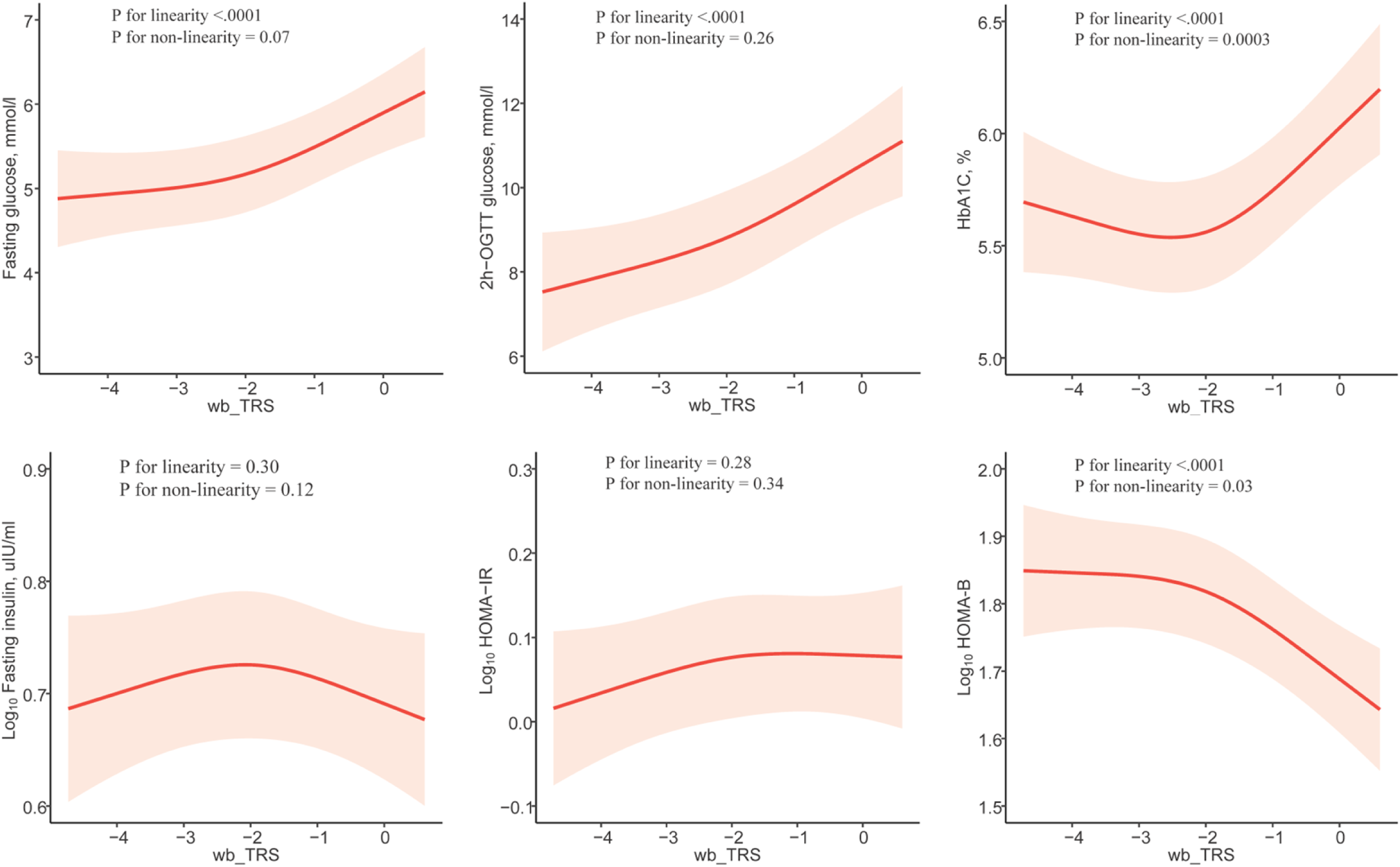
Restricted cubic spline analysis for associations between the wb-TRS and cardiometabolic traits in testing dataset. Data were adjusted for age, sex, body mass index, family history of diabetes, waist circumference, smoking and drinking status, education level, physical activity, systolic and diastolic blood pressure, triglycerides, low-density lipoprotein cholesterol and anti-diabetic therapy.

**Table 2.** Association of each 1-unit increment in the wb_TRS with cardiometabolic traits (n=1105) Data were β and standard error (SE). Liner regression model was used to evaluate associations of the wb_TRS (independent) with cardiometabolic traits (dependent variables). Model 1, adjusted for age, sex, body mass index, and family history of diabetes; Model 2, further adjusted for waist circumference, smoking and drinking status, education level, physical activity, systolic and diastolic blood pressure, triglycerides, LDL-C and anti-diabetic therapy. Abbreviation: HDL-C, high-density lipoprotein cholesterol; LDL-C, low-density lipoprotein cholesterol; OGTT, oral glucose tolerance test; HbA1c, glycated hemoglobin A1c. HOMA-IR, homeostasis model assessment of insulin resistance; and HOMA-β, homeostasis model assessment of β cell function.

|  | Training dataset |  | Testing dataset |  |
| --- | --- | --- | --- | --- |
| | $\beta$ (SE) | *P* value | $\beta$ (SE) | *P* value |
| Body mass index, kg/m<sup>2</sup> |  |  |  |  |
| Model 1 | 0.09 (0.06) | 0.12 | 0.08 (0.10) | 0.43 |
| Model 2 | -0.05 (0.04) | 0.22 | -0.003 (0.08) | 0.97 |
| Waist circumference, cm |  |  |  |  |
| Model 1 | 0.38 (0.14) | 0.01 | 0.14 (0.23) | 0.53 |
| Model 2 | 0.35 (0.14) | 0.01 | 0.20 (0.24) | 0.41 |
| Systolic blood pressure, mmHg |  |  |  |  |
| Model 1 | 1.52 (0.49) | 0.002 | 0.80 (0.82) | 0.33 |
| Model 2 | 1.00 (0.39) | 0.01 | 0.45 (0.65) | 0.49 |
| Diastolic blood pressure, mmHg |  |  |  |  |
| Model 1 | 0.41 (0.28) | 0.14 | 0.02 (0.47) | 0.97 |
| Model 2 | -0.07 (0.22) | 0.77 | -0.35 (0.37) | 0.36 |
| Total cholesterol, mmol/l |  |  |  |  |
| Model 1 | -0.06 (0.03) | 0.03 | 0.11 (0.05) | 0.04 |
| Model 2 | -0.01 (0.01) | 0.22 | -0.002 (0.01) | 0.86 |
| Triglycerides, mmol/l |  |  |  |  |
| Model 1 | 0.09 (0.03) | 0.001 | 0.06 (0.03) | 0.08 |
| Model 2 | 0.09 (0.03) | 0.001 | 0.04 (0.03) | 0.29 |
| HDL-C, mmol/l |  |  |  |  |
| Model 1 | -0.02 (0.01) | 0.01 | 0.02 (0.01) | 0.25 |
| Model 2 | -0.01 (0.01) | 0.43 | 0.01 (0.01) | 0.43 |
| LDL-C, mmol/l |  |  |  |  |
| Model 1 | -0.04 (0.02) | 0.03 | 0.08 (0.04) | 0.04 |
| Model 2 | -0.06 (0.02) | 0.01 | 0.06 (0.04) | 0.12 |
| Fasting glucose, mmol/l |  |  |  |  |
| Model 1 | 0.42 (0.03) | <.0001 | 0.34 (0.06) | <.0001 |
| Model 2 | 0.25 (0.03) | <.0001 | 0.23 (0.06) | <.0001 |
| 2h-OGTT glucose, mmol/l |  |  |  |  |
| Model 1 | 1.16 (0.08) | <.0001 | 0.84 (0.13) | <.0001 |
| Model 2 | 1.01 (0.08) | <.0001 | 0.68 (0.14) | <.0001 |
| HbA1c, % |  |  |  |  |
| Model 1 | 0.21 (0.02) | <.0001 | 0.19 (0.04) | <.0001 |
| Model 2 | 0.07 (0.02) | <.0001 | 0.10 (0.03) | 0.002 |
| Log Fasting insulin, uIU/ml |  |  |  |  |
| Model 1 | 0.03 (0.005) | <.0001 | 0.0003 (0.01) | 0.97 |
| Model 2 | 0.01 (0.01) | 0.02 | -0.004 (0.01) | 0.61 |
| Log HOMA-IR |  |  |  |  |
| Model 1 | 0.05 (0.01) | <.0001 | 0.02 (0.01) | 0.02 |
| Model 2 | 0.03 (0.01) | <.0001 | 0.01 (0.01) | 0.24 |
| Log HOMA-β | | | | |
| Model 1 | -0.04 (0.01) | <.0001 | -0.05 (0.01) | <.0001 |
| Model 2 | -0.03 (0.01) | <.0001 | -0.04 (0.01) | <.0001 |

The GEE analysis by using the 3 repeated measurements of glucose metabolism traits and wb-TRS at baseline, 3 month- and 6 month-visits during the intervention showed that each 1-unit increment in wb-TRS was associated with 0.11mmol/l (SE 0.05) and 0.15mmol/l (SE 0.02) increment of 1h- and 2h-OGTT glucose, respectively. Interestingly, the baseline wb-TRS significantly affected the dynamic changes of cardiometabolic traits at 6-months of dietary intervention **(Table 3)**. Baseline wb-TRS significantly associated with changes in average daytime, nighttime and 24h glucose and HbA1c levels and the AUC of glucose measured in the continuous glucose monitoring at 6 month of diet intervention (all P<0.03).

**Table 3.** The effects of baseline wb-TRS on changes of glucose metabolic traits during the 6-month dietary intervention (n=253) Data were change (β) and standard error (SE). P values were from the general linear models after adjusted for sex, baseline age, body mass index, waist circumference, smoking and drinking status, education level, physical activity, systolic and diastolic blood pressure, triglycerides, LDL-C and baseline values for the respective outcome trait. Change (Δ) level was calculated as the numeric value at follow-up visits minus that at baseline. Estimated HbA1c was calculated using the formula: (average glucose+46.7)/28.7.

|  | At 3 months |  | At 6 months |  |
| --- | --- | --- | --- | --- |
| Glucose metabolic traits | $\beta$ (SE) | P value | $\beta$ (SE) | P value |
| OGTT |  |  |  |  |
| ΔFasting glucose, mmol/l | -0.01 (0.02) | 0.72 | 0.01 (0.02) | 0.64 |
| Δ0.5h-OGTT glucose, mmol/l | 0.03 (0.05) | 0.51 | -0.0005 (0.06) | 0.99 |
| Δ1h-OGTT glucose, mmol/l | 0.03 (0.08) | 0.74 | 0.12 (0.08) | 0.15 |
| Δ2h-OGTT glucose, mmol/l | 0.04 (0.07) | 0.59 | 0.10 (0.07) | 0.16 |
| Continuous glucose monitoring (CGM) |  |  |  |  |
| ΔAverage glucose, mmol/l | -0.21 (0.45) | 0.64 | 1.42 (0.55) | 0.01 |
| ΔAverage daytime glucose, mg/dl | -0.26 (0.48) | 0.59 | 1.55 (0.60) | 0.01 |
| ΔAverage nighttime glucose, mg/dl | -0.16 (0.42) | 0.71 | 1.09 (0.48) | 0.03 |
| ΔAUC glucose, mmol*h/l | 0.21 (0.45) | 0.65 | 1.41 (0.55) | 0.01 |
| ΔAUC daytime glucose, mmol*h/l | 0.26 (0.48) | 0.59 | 1.56 (0.60) | 0.01 |
| ΔAUC nighttime glucose, mmol*h/l | 0.16 (0.42) | 0.70 | 0.21 (0.45) | 0.65 |
| ΔEstimated HbA1c | -0.004 (0.02) | 0.81 | 0.05 (0.02) | 0.01 |
| ΔEstimated daytime HbA1c | -0.01 (0.02) | 0.55 | 0.06 (0.02) | 0.01 |
| ΔEstimated nighttime HbA1c | 0.01 (0.01) | 0.58 | 0.04 (0.02) | 0.03 |

### wb-TRS performed well in predicting T2D

In both training and testing dataset, wb-TRS performed well in predicting nonobese T2D, even when adding wb-TRS into a model with fasting glucose, triglycerides, and the primary model (**Fig 3**). In total dataset, a model with wb-TRS along with fasting glucose, triglycerides, and variables in primary model significantly improved the AUC (0.93), as compared to the primary model (AUC =0.65), the fasting glucose and triglycerides model (AUC=0.82), and the fasting glucose, triglycerides adding to the primary model (full model, AUC =0.84) for predicting T2D (**Table S3**). The wb-TRS improved predicting performance in a model with fasting glucose, triglycerides, and the demographic and anthropometry parameters (AUC-TRS+full: 0.93 [0.91-0.95] vs. AUC-full: 0.84 [95% CI 0.81-0.87]; P < 0.0001).

**Fig 3.**
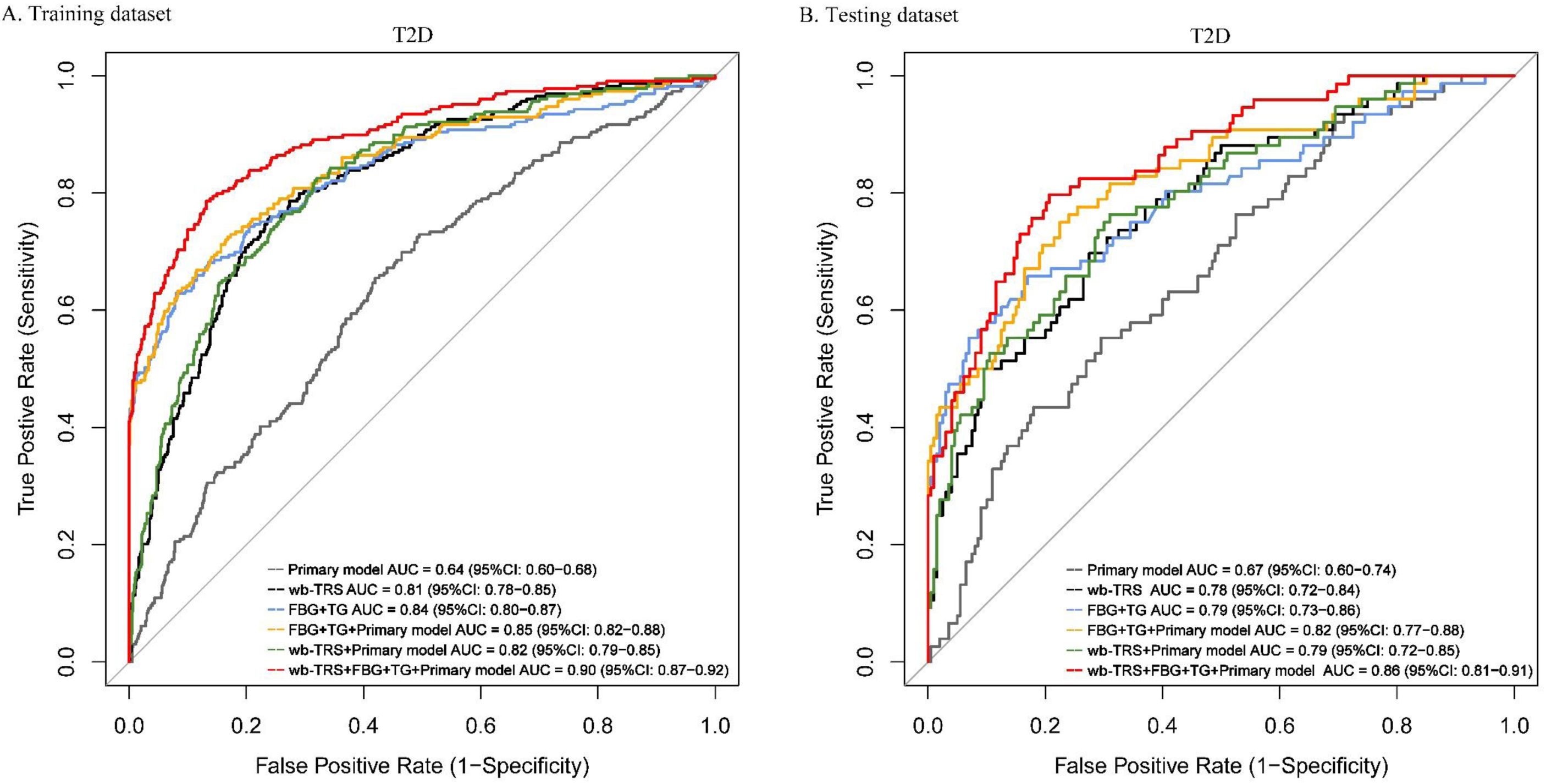
Receiver operator characteristic curves for prediction of type 2 diabetes. Primary model comprised age, sex, BMI and family history of diabetes. Abbreviations: wb-TRS, whole blood-based transcriptional risk score; BMI, body mass index; FBG, fasting glucose index; TG, triglycerides; and AUC, area under the curve.

### The biological pathway and mechanism analysis

The Mitch analysis compared the bottom versus middle and top versus middle tertiles of wb-TRS (**Fig 4**). The top versus middle tertile group comparison demonstrated downregulation of MYC target V2, and the bottom versus middle tertile group comparison demonstrated upregulation of oxidative phosphorylation, mTORC1 signaling, cholesterol metabolism, peroxisome, fatty acid metabolism, etc. **(Fig 4A)**. The significant metabolic pathways were displayed in **Fig 4B.** The small GTPase (RAB1A gene) enriched in mTORC1 signaling, fatty acid oxidation gene (CPT1A) enriched in oxidative phosphorylation pathway, and cholesterol efflux gene (ABCA1) enriched in cholesterol metabolism pathway.

**Fig 4.**
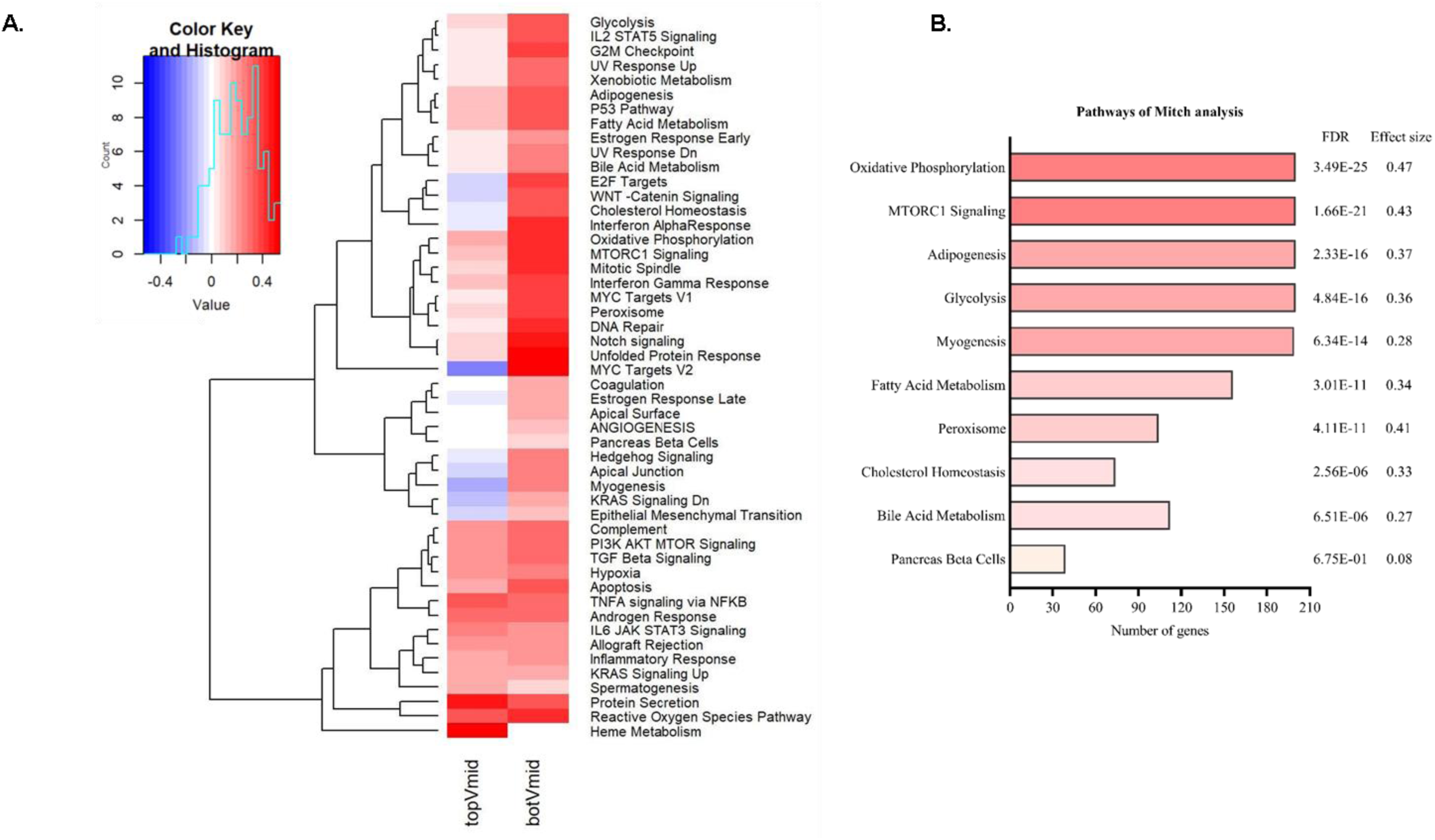
Mitch analysis. The gene expression level in the top and bottom tertile of the wb-TRS were compared with those in the middle tertile. **A**, a heatmap of enrichment scores for gene expression contrasts between these two comparisons with hierarchical clustering. Cells in red indicate a positive enrichment score, whereas blue indicated a negative enrichment score. **B**, the significant metabolic pathways. Abbreviations: botVmid, bottom versus middle tertile; topVmid, top versus middle tertile.

The Reactome analyses suggested that the 144 gene transcripts included in wb-TRS were involved in NR1H2 (LXRβ) and NR1H3 (LXRα) regulate gene expression to limit cholesterol uptake, regulation of lipid metabolism by PPAR alpha pathway, etc. The cholesterol uptake suppression gene (MYLIP) enriched in LXRβ and LXRα regulate gene expression to limit cholesterol uptake pathway (**Table S4**).

### Causal relationships between gene transcripts included in wb_TRS and T2D

Gene transcripts (n=19) with no validated IVs were removed from TWMR analysis. Among the 125 remaining gene transcripts, 6 gene transcripts (GCC1-PAX4, AKR1A1, AAMP, RAB1A, ITGA1, and ZNF639) were causally linked to T2D risk in AGEN and 1 gene transcript (RAB1A) was successfully validated in DIAGRAM (**Table S5**, P < 0.0004 =0.05/125, where 125 corresponds to the number of gene transcripts tested in TWMR). Specifically, RAB1A enriched in mTOCR1 signaling is causally associated with 38% (95% CI: 1.18-1.62, P=8.0×10^-5^) and 47% (95% CI: 1.21-1.77, P=5.4×10^-5^) increase of T2D risk in AGEN and DIAGRAM, respectively. GCC1-RAX4 converted 17% decrease of T2D risk (95% CI: 0.80-0.88, P=5.2×10^-15^) in AGEN.

## Discussion

In this study, we developed a wb-TRS for T2D risk in a longitudinal community-based cohort in which the participants were middle aged and elderly and maintained nonobese for up to 10 years. The wb-TRS was positively associated with risk of T2D and could reflect dynamic change in glucose metabolism such as fasting and 2h-post loading glucose, HbA1c, and beta cell dysfunction in the observational cohort. Furthermore, the baseline wb-TRS well associated with the changes of glucose metabolism measured in the continuous glucose monitoring in response to the 6-month dietary intervention in a panel of younger and overweight participants. The wb-TRS performed better and was a complementarity to fasting glucose, triglycerides and the demographic and anthropometric factors in predicting T2D. The wb-TRS supported several gene transcripts (RAB1A and GCC1-RAX4) and biological pathways (oxidative phosphorylation, cholesterol metabolism, and mTORC1 signaling) that might involve in the etiology of nonobese T2D.

To the best of our knowledge, the present study is the first to examine the effects of wb-TRS on nonobese T2D risk and dynamic change of related cardiometabolic traits by using peripheral blood transcriptional markers. Recently, several studies proposed the concept of TRS or polygenetic TRS, which used cumulative gene expression information to provide an additional layer of biological interpretability and performed better than both traditional risk factors and polygenic risk score for predicting Crohn’s disease, chronic obstructive pulmonary disease, and lung function [8, 26–28]. Our study is in line with previous studies showing transcriptional variance is a reliable intermediary mediating associations of genotypes with complex phenotypes [5, 6, 29] and provides evidence of implement of blood transcriptional profile could yield a promising better predictive effect on T2D and the dynamic changes in related cardiometabolic traits.

In our study, the wb-TRS could well predict dynamic changes in glucose metabolism in response to 6 months of 25% calorie restriction dietary intervention in overweight/obese subjects [23]. In our trial (DPMH), a 25% calorie restriction produced an appreciable weight reduction and improved glucose homeostasis [23]. Calorie restriction improves cardiometabolic risk including atherogenic dyslipidemia (elevated triglycerides and fasting glucose, reduced HDL-C, diabetes, hypertension, and large waist circumference) [30, 31]. Previous findings from the Comprehensive Assessment of Long-term Effects of Reducing Intake of Energy (CALERIETM) study, a 24-month, multicenter, randomized controlled calorie restriction trial, showed that in young to middle aged, normal to slightly overweight adults, two years of >12% caloric restriction improved traditional cardiometabolic risk factors within ranges considered normal [31]. Recent study suggested that in healthy adults without obesity, a 12% calorie restriction led to improved reducing inflammation and branched chain amino acids and shifting lipoproteins from atherogenic to cholesterol transporting [32]. We suspected that the genes and pathways included in the wb-TRS played key roles in cardiometabolic beneficial effects in response to the calorie restriction dietary intervention, which might help to explore the underlying mechanisms for the beneficial effects.

Our results indicated several potential pathways and gene transcripts that might involve in the etiology process of nonobese T2D, which suggested that transcriptional information in blood could be capable of reflecting diseases risk and could bridge the knowledge gap on how gene expression affects nonobese T2D. We found mTORC1 signaling directly regulates β-cell functional maturation and identity maintenance [33]. Of note, by using TWMR analysis, we also demonstrated that the RAB1A gene encoding a small GTPase, which is enriched in mTORC1 signaling, was positively and causally associated with T2D risk in both east Asians and Europeans. Another T2D susceptibily gene (GCC1-PAX4) was also causally associated with T2D risk in east Asians [34]. PAX4 implicated in a Japanese case of maturity onset diabetes of the young that is only 30kb further away from GCC1 is an outstanding candidate for T2D given its involvement in pancreatic islet development [34]. This molecular mechanism of nonobese T2D was consistent with that the nonobese Asian Indians with young-onset diabetes have lower genetically determined beta cell function [35]. We identified oxidative phosphorylation and cholesterol metabolism pathways involved in nonobese T2D. Of note, fatty acid oxidation gene (CPT1A) enriched in oxidative phosphorylation pathway might mediate mitochondrial overload increasing insulin resistance in muscle, lipid accumulation in liver, and lipotoxicity in pancreatic beta cells thus involved in homeostasis of glucose metabolism [36, 37]. Cholesterol efflux gene (ABCA1) enriched in cholesterol metabolism pathway might via its downregulation to inhibit cholesterol transportion to HDL and excretion to bile and faeces, and then increase lipids profile and deteriorates cardio-metabolism [38].

In Reactome analysis, cholesterol uptake suppression gene (MYLIP) enriched in LXRβ and LXRα regulate gene expression to limit cholesterol uptake pathway. Inhibition of MYLIP increases cellular cholesterol accumulation and leads to pancreatic β-cell dysfunction and insulin resistance [39]. We suspected that cholesterol metabolism in liver and pancreas and beta cell dysfunction involved in nonobese T2D but warrant further study in the future. Besides, the consistency of transcriptional levels of these genes in whole-blood and specific tissue is another interesting issue.

The major strengths of our current study lie in our adopted analytical approaches as well as the well-characterized study cohorts. We adopted high-throughput RNA-seq technology, a penalized regression model with 10-fold cross validation method to develop the wb-TRS, and successfully validated its performance in an independent external cohort, which guaranteed the statistical power and robustness of our findings and conclusions. In terms of the study cohorts, our initial cohort included T2D whose BMI maintained normal for up to 10 years, which are expected to better capture underling pathophysiology mechanism of nonobese T2D.

Our study, however, also has several limitations. First, transcriptome data were measured only at 10 years follow-up visit in initial cohort, which could not establish a causal relationship between the wb-TRS and nonobese T2D risk. It would also be greater interesting to see whether transcriptional change would continue to be associated with incidence risk of nonobese T2D. Second, complex diseases are final phenotypes of transcriptional dysregulation in multiple tissues. Though an appreciable proportion of genes expressed in tissues is expressed in peripheral whole blood [40], it still should be better to interpret the present wb-TRS as one of potential essential complementary prediction tools for nonobese T2D, rather than presenting an exact pathophysiological process of it. Nevertheless, the individual’s blood transcriptome performed well in predicting tissue-specific expression and performed as well as measured tissue expression in predicting complex diseases [5]. Third, we did not incorporate metabolic markers in the analysis, which restrained us from comparing the predictive effect between them. Jointly considering the genetic and metabolic profiling and transcriptional level of alterations would provide more precise and accurate evaluation of T2D risk and prediction efficacy. Last, both the initial and vailidation cohort consisted of participants from east Asia, cautions should be exercised to generalize the finding to other ethnicity groups.

In conclusion, we developed a wb-TRS that comprising a panel of genes based on their blood expression levels, and that may involve important biological function pathways was positively associated with nonobese T2D risk and predicted dynamic change in glucose metabolic traits. The study supported that blood transcriptome highly related to and reflect impaired glucose metabolism. It may lend insight into discerning potential causal genes and biological mechanisms in the pathogenesis of nonobese T2D.

## Data Availability

The eQTLs data used in this study are available on the eQLTGen Consortium website [https://eqtlgen.org/]. The summary statistics of genome-wide association study of T2D was available in the Asian Genetic Epidemiology Network (AGEN) [https://blog.nus.edu.sg/agen/] and the DIAbetes Genetics Replication And Meta analysis (DIAGRAM) [http://www.diagram-consortium.org/downloads.html/]. RNA seq related additional data reported in this manuscript are available from the corresponding author upon reasonable request.

## Abbreviations

AUC: area under the curve
BMI: body mass index
CPM: count per million
CI: confidence interval
DBP: diastolic blood pressure
DPMH: Dietary Pattern and Metabolic Health
eQTL: expression quantitative trait locus
GEE: generalized estimating equation
HbA1c: hemoglobin A1c
HOMA-IR: homeostasis model assessment of insulin resistance
HOMA-β: homeostasis model assessment of beta cell function
HDL-C: high-density lipoprotein cholesterol
MET: metabolic equivalent of task
Mitch analysis: a multi-contrast gene set enrichment analysis
LDL-C: low-density lipoprotein cholesterol
LASSO: least absolute shrinkage and selection operator
OGTT: oral glucose tolerance test
OR: odds ratio
RCS: restricted cubic spline
RNA-seq: RNA sequencing
SBP: systolic blood pressure
T2D: Type 2 diabetes
TRS: transcriptional risk score
TWMR: Transcriptome-wide Mendelian randomization
TSGE: Tissue-specific gene expression

## Acknowledgments

We thank all the study participants for their participation and contribution.

## Data availability

The eQTLs data used in this study are available on the eQLTGen Consortium website [https://eqtlgen.org/]. The summary statistics of genome-wide association study of T2D was available in the Asian Genetic Epidemiology Network (AGEN) [https://blog.nus.edu.sg/agen/] and the DIAbetes Genetics Replication And Meta-analysis (DIAGRAM) [http://www.diagram-consortium.org/downloads.html/]. RNA-seq related additional data reported in this manuscript are available from the corresponding author upon reasonable request.

## Contribution statement

M.X., Y.B., and J.W. contributed to the conception and design of the study. Y.H., H.D., and N.C. performed statistical analysis and interpreted data. Y.H., and M.X. drafted the manuscript. G.N. and W.W. revised the manuscript critically for important intellectual content. Z.Z., Q.W., T.H., J.Z., T.W., M.L., H.L., S.W., R.Z., J.L., Y.X., Y.C., and R.L. contributed to the acquisition of data. All authors were involving in revising the manuscript and had final approval of the version to be published. M.X., Y.B. and J.W. are the guarantors of this work and, as such, had full access to all the data in the study and take responsibility for the integrity of the data and the accuracy of the data analysis.

## Supporting information

**Table S1.** Clinical characteristics of study participants.

**Table S2.** Weights of 144 gene transcripts included in wb-TRS, ordered from most negative to most positive associated with nonobese type 2 diabetes.

**Table S3.** Receiver operator characteristic curves for prediction of nonobese type 2 diabetes.

**Table S4.** Top30 Reactome pathways associated with genes in wb-TRS.

**Table S5.** Transcriptome wide Mendelian randomization shows the causal effect of gene transcripts included in wb_TRS on type 2 diabetes.

**Fig S1.** Flowchart of study participants.

**Fig S2.** Cross-validation results for least absolute shrinkage and selection operator (LASSO) logistics-model for nonobese type 2 diabetes in training dataset.

**Fig S3.** Violin plot of wb-TRS and gene transcripts most negatively (FUOM) and positively (RPF1) associated with nonobese type 2 diabetes.

**Fig S4.** Stratified analysis. Data are odds ratio (OR) and 95% confidence interval (CI) calculated using multivariable logistic regression models. The results were adjusted for age, sex, body mass index, family history of diabetes, waist circumference, smoking and drinking status, education level, physical activity, systolic and diastolic blood pressure, total cholesterol, triglycerides, high-density and low-density lipoprotein cholesterol.

## References

1. IDF Diabetes Atlas 2021 – 10th edition. www.diabetesatlas.org.

2. Hahn SJ, Kim S, Choi YS, Lee J, Kang J. Prediction of type 2 diabetes using genome-wide polygenic risk score and metabolic profiles: A machine learning analysis of population-based 10-year prospective cohort study. EBioMedicine. 2022;86:104383. Epub 20221130. doi: 10.1016/j.ebiom.2022.104383. PubMed PMID: 36462406; PubMed Central PMCID: PMCPMC9713286.

3. Wang S, Li M, Lin H, Wang G, Xu Y, Zhao X, et al. Amino acids, microbiota-related metabolites, and the risk of incident diabetes among normoglycemic Chinese adults: Findings from the 4C study. Cell Rep Med. 2022;3(9):100727. Epub 20220822. doi: 10.1016/j.xcrm.2022.100727. PubMed PMID: 35998626; PubMed Central PMCID: PMCPMC9512668.

4. Padilla-Martinez F, Wojciechowska G, Szczerbinski L, Kretowski A. Circulating Nucleic Acid-Based Biomarkers of Type 2 Diabetes. Int J Mol Sci. 2021;23(1). Epub 20211228. doi: 10.3390/ijms23010295. PubMed PMID: 35008723; PubMed Central PMCID: PMCPMC8745431.

5. Mahashweta B, Kun W, Eytan R, Sridhar H. Predicting tissue-specific gene expression from whole blood transcriptome. 2021;7:eabd6991.

6. Yao S, Zhang X, Zou SC, Zhu Y, Li B, Kuang WP, et al. A transcriptome-wide association study identifies susceptibility genes for Parkinson’s disease. NPJ Parkinsons Dis. 2021;7(1):79. Epub 20210909. doi: 10.1038/s41531-021-00221-7. PubMed PMID: 34504106; PubMed Central PMCID: PMCPMC8429416.

7. Wang B, Lunetta KL, Dupuis J, Lubitz SA, Trinquart L, Yao L, et al. Integrative Omics Approach to Identifying Genes Associated With Atrial Fibrillation. Circ Res. 2020;126(3):350–60. Epub 20191205. doi: 10.1161/CIRCRESAHA.119.315179. PubMed PMID: 31801406; PubMed Central PMCID: PMCPMC7004281.

8. Moll M, Boueiz A, Ghosh AJ, Saferali A, Lee S, Xu Z, et al. Development of a Blood-based Transcriptional Risk Score for Chronic Obstructive Pulmonary Disease. Am J Respir Crit Care Med. 2022;205(2):161–70. doi: 10.1164/rccm.202107-1584OC. PubMed PMID: 34739356; PubMed Central PMCID: PMCPMC8787248.

9. Li X, Su X, Liu J, Li H, Li M, andMe Research T, et al. Transcriptome-wide association study identifies new susceptibility genes and pathways for depression. Transl Psychiatry. 2021;11(1):306. Epub 20210521. doi: 10.1038/s41398-021-01411-w. PubMed PMID: 34021117; PubMed Central PMCID: PMCPMC8140098.

10. Teufel F, Seiglie JA, Geldsetzer P, Theilmann M, Marcus ME, Ebert C, et al. Body-mass index and diabetes risk in 57 low-income and middle-income countries: a cross-sectional study of nationally representative, individual-level data in 685 616 adults. The Lancet. 2021;398(10296):238–48. doi: 10.1016/s0140-6736(21)00844-8.

11. Taylor R, Holman RR. Normal weight individuals who develop type 2 diabetes: the personal fat threshold. Clin Sci (Lond). 2015;128(7):405–10. doi: 10.1042/CS20140553. PubMed PMID: 25515001.

12. Zou X, Zhou X, Zhu Z, Ji L. Novel subgroups of patients with adult-onset diabetes in Chinese and US populations. The Lancet Diabetes & Endocrinology. 2019;7(1):9–11. doi: 10.1016/s2213-8587(18)30316-4.

13. Zaharia OP, Strassburger K, Strom A, Bönhof GJ, Karusheva Y, Antoniou S, et al. Risk of diabetes-associated diseases in subgroups of patients with recent-onset diabetes: a 5-year follow-up study. The Lancet Diabetes & Endocrinology. 2019;7(9):684–94. doi: 10.1016/s2213-8587(19)30187-1.

14. Ahlqvist E, Storm P, Käräjämäki A, Martinell M, Dorkhan M, Carlsson A, et al. Novel subgroups of adult-onset diabetes and their association with outcomes: a data-driven cluster analysis of six variables. The Lancet Diabetes & Endocrinology. 2018;6(5):361–9. doi: 10.1016/s2213-8587(18)30051-2.

15. Wesolowska-Andersen A, Brorsson CA, Bizzotto R, Mari A, Tura A, Koivula R, et al. Four groups of type 2 diabetes contribute to the etiological and clinical heterogeneity in newly diagnosed individuals: An IMI DIRECT study. Cell Rep Med. 2022;3(1):100477. Epub 20220104. doi: 10.1016/j.xcrm.2021.100477. PubMed PMID: 35106505; PubMed Central PMCID: PMCPMC8784706.

16. Spracklen CN, Horikoshi M, Kim YJ, Lin K, Bragg F, Moon S, et al. Identification of type 2 diabetes loci in 433,540 East Asian individuals. Nature. 2020;582(7811):240–5. Epub 20200506. doi: 10.1038/s41586-020-2263-3. PubMed PMID: 32499647; PubMed Central PMCID: PMCPMC7292783.

17. Wang B, Li M, Zhao Z, Lu J, Chen Y, Xu Y, et al. Urinary bisphenol A concentration and glucose homeostasis in non-diabetic adults: a repeated-measures, longitudinal study. Diabetologia. 2019;62(9):1591–600. doi: 10.1007/s00125-019-4898-x.

18. Hao M, Ding L, Xuan L, Wang T, Li M, Zhao Z, et al. Urinary bisphenol A concentration and the risk of central obesity in Chinese adults: A prospective study. J Diabetes. 2018;10(6):442–8. Epub 20170329. doi: 10.1111/1753-0407.12531. PubMed PMID: 28097815.

19. Ning G, Bi Y, Wang T, Xu M, Xu Y, Huang Y, et al. Relationship of Urinary Bisphenol A Concentration to Risk for Prevalent Type 2 Diabetes in Chinese Adults. Annals of Internal Medicine. 2011 Sep 20;155(6):368–74.

20. American Diabetes A. Diagnosis and classification of diabetes mellitus. Diabetes Care. 2010;33 Suppl 1:S62–9. doi: 10.2337/dc10-S062. PubMed PMID: 20042775; PubMed Central PMCID: PMCPMC2797383.

21. Juergen C, Matthias M. 1D and 2D annotation enrichment: a statistical method integrating quantitative proteomics with complementary high-throughput data. BMC Bioinformatics. 2012;13(Suppl 16):S12.

22. Fabregat A, Sidiropoulos K, Viteri G, Forner O, Marin-Garcia P, Arnau V, et al. Reactome pathway analysis: a high-performance in-memory approach. BMC Bioinformatics. 2017;18(1):142. Epub 20170302. doi: 10.1186/s12859-017-1559-2. PubMed PMID: 28249561; PubMed Central PMCID: PMCPMC5333408.

23. Luo Y, Wang J, Sun L, Gu W, Zong G, Song B, et al. Isocaloric-restricted Mediterranean Diet and Chinese Diets High or Low in Plants in Adults With Prediabetes. J Clin Endocrinol Metab. 2022;107(8):2216–27. doi: 10.1210/clinem/dgac303. PubMed PMID: 35579171; PubMed Central PMCID: PMCPMC9282247.

24. Sladek R, Rocheleau G, Rung J, Dina C, Shen L, Serre D, et al. A genome-wide association study identifies novel risk loci for type 2 diabetes. Nature. 2007;445(7130):881-5. Epub 20070211. doi: 10.1038/nature05616. PubMed PMID: 17293876.

25. Porcu E, Rueger S, Lepik K, e QC, Consortium B, Santoni FA, et al. Mendelian randomization integrating GWAS and eQTL data reveals genetic determinants of complex and clinical traits. Nat Commun. 2019;10(1):3300. Epub 20190724. doi: 10.1038/s41467-019-10936-0. PubMed PMID: 31341166; PubMed Central PMCID: PMCPMC6656778.

26. Hu X, Qiao D, Kim W, Moll M, Balte PP, Lange LA, et al. Polygenic transcriptome risk scores for COPD and lung function improve cross-ethnic portability of prediction in the NHLBI TOPMed program. Am J Hum Genet. 2022;109(5):857–70. Epub 20220405. doi: 10.1016/j.ajhg.2022.03.007. PubMed PMID: 35385699; PubMed Central PMCID: PMCPMC9118106.

27. Liang Y, Pividori M, Manichaikul A, Palmer AA, Cox NJ, Wheeler HE, et al. Polygenic transcriptome risk scores (PTRS) can improve portability of polygenic risk scores across ancestries. Genome Biol. 2022;23(1):23. Epub 20220113. doi: 10.1186/s13059-021-02591-w. PubMed PMID: 35027082; PubMed Central PMCID: PMCPMC8759285.

28. Marigorta UM, Denson LA, Hyams JS, Mondal K, Prince J, Walters TD, et al. Transcriptional risk scores link GWAS to eQTLs and predict complications in Crohn’s disease. Nat Genet. 2017;49(10):1517–21. Epub 20170814. doi: 10.1038/ng.3936. PubMed PMID: 28805827; PubMed Central PMCID: PMCPMC5745037.

29. Nica AC, Montgomery SB, Dimas AS, Stranger BE, Beazley C, Barroso I, et al. Candidate causal regulatory effects by integration of expression QTLs with complex trait genetic associations. PLoS Genet. 2010;6(4):e1000895. Epub 20100401. doi: 10.1371/journal.pgen.1000895. PubMed PMID: 20369022; PubMed Central PMCID: PMCPMC2848550.

30. Luigi F, Timothy E MS, Klein., John O H. Long-term calorie restriction is highly effective in reducing the risk for atherosclerosis in humans. Proc Natl Acad Sci USA. 2004;101 (17):6659–63.

31. Kraus WE, Bhapkar M, Huffman KM, Pieper CF, Krupa Das S, Redman LM, et al. 2 years of calorie restriction and cardiometabolic risk (CALERIE): exploratory outcomes of a multicentre, phase 2, randomised controlled trial. The Lancet Diabetes & Endocrinology. 2019;7(9):673–83. doi: 10.1016/s2213-8587(19)30151-2.

32. Huffman KM, Parker DC, Bhapkar M, Racette SB, Martin CK, Redman LM, et al. Calorie restriction improves lipid-related emerging cardiometabolic risk factors in healthy adults without obesity: Distinct influences of BMI and sex from CALERIE a multicentre, phase 2, randomised controlled trial. EClinicalMedicine. 2022;43:101261. Epub 20220103. doi: 10.1016/j.eclinm.2021.101261. PubMed PMID: 35028547; PubMed Central PMCID: PMCPMC8741476.

33. Ni Q, Sun J, Wang Y, Wang Y, Liu J, Ning G, et al. mTORC1 is required for epigenetic silencing during beta-cell functional maturation. Mol Metab. 2022;64:101559. Epub 20220805. doi: 10.1016/j.molmet.2022.101559. PubMed PMID: 35940555; PubMed Central PMCID: PMCPMC9418906.

34. Cho YS, Chen CH, Hu C, Long J, Ong RT, Sim X, et al. Meta-analysis of genome-wide association studies identifies eight new loci for type 2 diabetes in east Asians. Nat Genet. 2011;44(1):67–72. Epub 20111211. doi: 10.1038/ng.1019. PubMed PMID: 22158537; PubMed Central PMCID: PMCPMC3582398.

35. Siddiqui MK, Anjana RM, Dawed AY, Martoeau C, Srinivasan S, Saravanan J, et al. Young-onset diabetes in Asian Indians is associated with lower measured and genetically determined beta cell function. Diabetologia. 2022;65(6):973–83. Epub 20220305. doi: 10.1007/s00125-022-05671-z. PubMed PMID: 35247066; PubMed Central PMCID: PMCPMC9076730.

36. Lai CQ, Parnell LD, Smith CE, Guo T, Sayols-Baixeras S, Aslibekyan S, et al. Carbohydrate and fat intake associated with risk of metabolic diseases through epigenetics of CPT1A. Am J Clin Nutr. 2020;112(5):1200–11. doi: 10.1093/ajcn/nqaa233. PubMed PMID: 32930325; PubMed Central PMCID: PMCPMC7657341.

37. Meng ZX, Yin Y, Lv JH, Sha M, Lin Y, Gao L, et al. Aberrant activation of liver X receptors impairs pancreatic beta cell function through upregulation of sterol regulatory element-binding protein 1c in mouse islets and rodent cell lines. Diabetologia. 2012;55(6):1733–44. Epub 20120314. doi: 10.1007/s00125-012-2516-2. PubMed PMID: 22415588.

38. Wang JQ, Li LL, Hu A, Deng G, Wei J, Li YF, et al. Inhibition of ASGR1 decreases lipid levels by promoting cholesterol excretion. Nature. 2022;608(7922):413–20. Epub 20220803. doi: 10.1038/s41586-022-05006-3. PubMed PMID: 35922515.

39. Hao M, Head WS, Gunawardana SC, Hasty AH, Piston DW. Direct effect of cholesterol on insulin secretion: a novel mechanism for pancreatic beta-cell dysfunction. Diabetes. 2007;56(9):2328–38. Epub 20070615. doi: 10.2337/db07-0056. PubMed PMID: 17575085.

40. Radzikowska U, Ding M, Tan G, Zhakparov D, Peng Y, Wawrzyniak P, et al. Distribution of ACE2, CD147, CD26, and other SARS-CoV-2 associated molecules in tissues and immune cells in health and in asthma, COPD, obesity, hypertension, and COVID-19 risk factors. Allergy. 2020;75(11):2829–45. Epub 20200824. doi: 10.1111/all.14429. PubMed PMID: 32496587; PubMed Central PMCID: PMCPMC7300910.

