## Supplemental figures and tables for "A whole blood-based transcriptional risk score for nonobese type 2 diabetes predicts dynamic changes in glucose metabolic traits"

**Table S1.** Clinical characteristics of study participants

| **Variable** | **Total samples** | **Training dataset** | **Testing dataset** | **P value** |
| --- | --- | --- | --- | --- |
| N (%) | 1105 | 829 (75) | 276 (25) | / |
| Women (%) | 737 (66.7) | 544 (65.6) | 193 (69.9) | 0.19 |
| Age, years | 64.5 ± 7.0 | 64.4 ± 7.1 | 64.8 ± 7.0 | 0.42 |
| High school or above, n (%) | 382 (34.6) | 298 (36.0) | 84 (30.4) | 0.09 |
| Physical activity, MET-h/week | 21.0 (7.0-29.4) | 21.0 (0-30.0) | 21.0 (3.0-28.0) | 0.51 |
| Current smoking, n (%) | 196 (17.8) | 156 (18.8) | 40 (14.5) | 0.10 |
| Current drinking, n (%) | 94 (8.5) | 77 (9.3) | 17 (6.2) | 0.11 |
| Family history of diabetes, n (%) | 189 (17.1) | 136 (16.4) | 53 (19.2) | 0.29 |
| Body mass index, kg/m2 | 22.6 ± 2.3 | 22.6 ± 2.3 | 22.6 ± 2.4 | 0.67 |
| Waist circumference, cm | 79.6 ± 7.9 | 79.9 ± 8.0 | 78.9 ± 7.6 | 0.08 |
| Systolic blood pressure, mmHg | 135.4 ± 19.9 | 135.6 ± 19.8 | 134.6 ± 20.2 | 0.44 |
| Diastolic blood pressure, mmHg | 75.6 ± 11.0 | 75.6 ± 10.9 | 75.5 ± 11.2 | 0.88 |
| Total cholesterol, mmol/l | 5.47 ± 1.10 | 5.46 ± 1.06 | 5.48 ± 1.21 | 0.77 |
| Triglycerides, mmol/l | 1.35 (1.00-1.88) | 1.35 (0.99-1.88) | 1.35 (1.02-1.87) | 0.55 |
| HDL-C, mmol/l | 1.43 ± 0.33 | 1.43 ± 0.34 | 1.42 ± 0.32 | 0.68 |
| LDL-C, mmol/l | 3.45 ± 0.80 | 3.44 ± 0.77 | 3.46 ± 0.86 | 0.73 |
| Fasting glucose, mmol/l | 5.67 ± 1.46 | 5.69 ± 1.47 | 5.61 ± 1.40 | 0.43 |
| 2h-OGTT glucose, mmol/l | 8.73 ± 3.37 | 8.76 ± 3.37 | 8.64 ± 3.36 | 0.63 |
| Fasting insulin, mIU/l | 6.10 (4.70-8.10) | 6.1 (4.8-8.2) | 6.0 (4.6-7.9) | 0.41 |
| 2h-OGTT insulin, mIU/l | 36.2 (23.2-57.8) | 35.7 (23.8-57.8) | 37.1 (21.5-58.7) | 0.84 |
| HbA1c, % | 5.82 ± 0.82 | 5.82 ± 0.80 | 5.87 ± 0.87 | 0.64 |
| HOMA-IR | 1.49 (1.09-2.07) | 1.52 (1.09-2.08) | 1.41 (1.08-2.01) | 0.32 |
| HOMA-β, % | 66.7 (48.9-91.8) | 66.7 (49.1-92.3) | 66.9 (47.9-90.3) | 0.69 |
| **Categorical variables** |  |  |  |  |
| Type 2 diabetes, n (%) | 305 (27.6) | 229 (27.6) | 76 (27.5) | 0.98 |
| Newly diagnosed type 2 diabetes, n (%) | 139 (12.6) | 105 (12.7) | 34 (12.3) | 0.89 |
| Type 2 diabetes with duration ≥5 years, n (%) | 158 (14.3) | 118 (14.2) | 40 (14.5) | 0.94 |
| Antidiabetic agents, n (%) | 106 (9.6) | 81 (9.8) | 25 (9.1) | 0.72 |

Data are presented as means ± standard deviation (SD), or medians (inter-quartile ranges) for skewed variables, or number (percent) for categorical variables. P values were calculated from one-way analysis of variance (ANOVA) for continuous variables and chi-square test for categorical variables. Abbreviations: HDL-C, high-density lipoprotein cholesterol; LDL-C, low-density lipoprotein cholesterol; OGTT, oral glucose tolerance test; HbA1c, glycated hemoglobin A1c; HOMA-IR, homeostasis model assessment of insulin resistance; and HOMA-β, homeostasis model assessment of β cell function.

**Table S2.** Weights of 144 gene transcripts included in wb-TRS, ordered from most negative to most positive associated with nonobese type 2 diabetes.

| **Gene_ID** | **Gene_name** | **Gene_type** | **Weight** |
| --- | --- | --- | --- |
| ENSG00000148803 | FUOM | protein_coding | -0.2498 |
| ENSG00000173599 | PC | protein_coding | -0.16115 |
| ENSG00000225528 | Z82206.1 | protein_coding | -0.11553 |
| ENSG00000149929 | HIRIP3 | protein_coding | -0.09632 |
| ENSG00000068394 | GPKOW | protein_coding | -0.08105 |
| ENSG00000160013 | PTGIR | protein_coding | -0.06879 |
| ENSG00000176974 | SHMT1 | protein_coding | -0.05567 |
| ENSG00000170291 | ELP5 | protein_coding | -0.05541 |
| ENSG00000273173 | SNURF | protein_coding | -0.05125 |
| ENSG00000075826 | SEC31B | protein_coding | -0.05 |
| ENSG00000196497 | IPO4 | protein_coding | -0.04978 |
| ENSG00000119711 | ALDH6A1 | protein_coding | -0.04419 |
| ENSG00000100211 | CBY1 | protein_coding | -0.04262 |
| ENSG00000106665 | CLIP2 | protein_coding | -0.04093 |
| ENSG00000174791 | RIN1 | protein_coding | -0.03848 |
| ENSG00000106991 | ENG | protein_coding | -0.03712 |
| ENSG00000167074 | TEF | protein_coding | -0.03554 |
| ENSG00000004478 | FKBP4 | protein_coding | -0.03355 |
| ENSG00000152465 | NMT2 | protein_coding | -0.03253 |
| ENSG00000003056 | M6PR | protein_coding | -0.03072 |
| ENSG00000117632 | STMN1 | protein_coding | -0.02728 |
| ENSG00000136463 | TACO1 | protein_coding | -0.02691 |
| ENSG00000198964 | SGMS1 | protein_coding | -0.02309 |
| ENSG00000084463 | WBP11 | protein_coding | -0.01974 |
| ENSG00000007944 | MYLIP | protein_coding | -0.01928 |
| ENSG00000157107 | FCHO2 | protein_coding | -0.01744 |
| ENSG00000044574 | HSPA5 | protein_coding | -0.01678 |
| ENSG00000063978 | RNF4 | protein_coding | -0.0141 |
| ENSG00000117448 | AKR1A1 | protein_coding | -0.01352 |
| ENSG00000165802 | NSMF | protein_coding | -0.01324 |
| ENSG00000091409 | ITGA6 | protein_coding | -0.01216 |
| ENSG00000141510 | TP53 | protein_coding | -0.01209 |
| ENSG00000248592 | STIMATE-MU | protein_coding | -0.01173 |
| ENSG00000140688 | RUSF1 | protein_coding | -0.01173 |
| ENSG00000100600 | LGMN | protein_coding | -0.01152 |
| ENSG00000143797 | MBOAT2 | protein_coding | -0.01132 |
| ENSG00000124813 | RUNX2 | protein_coding | -0.01029 |
| ENSG00000135049 | AGTPBP1 | protein_coding | -0.00964 |
| ENSG00000164080 | RAD54L2 | protein_coding | -0.00964 |
| ENSG00000130653 | PNPLA7 | protein_coding | -0.00941 |
| ENSG00000172725 | CORO1B | protein_coding | -0.00877 |
| ENSG00000206047 | DEFA1 | protein_coding | -0.00813 |
| ENSG00000178732 | GP5 | protein_coding | -0.00718 |
| ENSG00000100147 | CCDC134 | protein_coding | -0.00669 |
| ENSG00000099337 | KCNK6 | protein_coding | -0.00647 |
| ENSG00000132823 | OSER1 | protein_coding | -0.00639 |
| ENSG00000204020 | LIPN | protein_coding | -0.00634 |
| ENSG00000166946 | CCNDBP1 | protein_coding | -0.00604 |
| ENSG00000151136 | BTBD11 | protein_coding | -0.00572 |
| ENSG00000178904 | DPY19L3 | protein_coding | -0.00569 |
| ENSG00000276900 | AC023157.2 | lncRNA | -0.00564 |
| ENSG00000198663 | C6orf89 | protein_coding | -0.00515 |
| ENSG00000214022 | REPIN1 | protein_coding | -0.00483 |
| ENSG00000069020 | MAST4 | protein_coding | -0.00436 |
| ENSG00000166527 | CLEC4D | protein_coding | -0.00428 |
| ENSG00000154358 | OBSCN | protein_coding | -0.00416 |
| ENSG00000114735 | HEMK1 | protein_coding | -0.00413 |
| ENSG00000116815 | CD58 | protein_coding | -0.00315 |
| ENSG00000071051 | NCK2 | protein_coding | -0.00195 |
| ENSG00000171456 | ASXL1 | protein_coding | -0.0019 |
| ENSG00000130066 | SAT1 | protein_coding | -0.00188 |
| ENSG00000275395 | FCGBP | protein_coding | -0.00188 |
| ENSG00000143546 | S100A8 | protein_coding | -0.00161 |
| ENSG00000106554 | CHCHD3 | protein_coding | -0.00118 |
| ENSG00000196565 | HBG2 | protein_coding | -0.00041 |
| ENSG00000179562 | GCC1-PAX4 | protein_coding | -0.00039 |
| ENSG00000116489 | CAPZA1 | protein_coding | -0.00021 |
| ENSG00000263264 | AC119396.1 | protein_coding | 0.000152 |
| ENSG00000101782 | RIOK3 | protein_coding | 0.000186 |
| ENSG00000105447 | GRWD1 | protein_coding | 0.000743 |
| ENSG00000130787 | HIP1R | protein_coding | 0.000892 |
| ENSG00000117643 | MAN1C1 | protein_coding | 0.001168 |
| ENSG00000112335 | SNX3 | protein_coding | 0.001276 |
| ENSG00000125863 | MKKS | protein_coding | 0.001328 |
| ENSG00000100324 | TAB1 | protein_coding | 0.001464 |
| ENSG00000100744 | GSKIP | protein_coding | 0.001765 |
| ENSG00000173868 | PHOSPHO1 | protein_coding | 0.001912 |
| ENSG00000168300 | PCMTD1 | protein_coding | 0.001991 |
| ENSG00000102908 | NFAT5 | protein_coding | 0.002274 |
| ENSG00000175061 | SNHG29 | lncRNA | 0.002289 |
| ENSG00000170345 | FOS | protein_coding | 0.002746 |
| ENSG00000148840 | PPRC1 | protein_coding | 0.002809 |
| ENSG00000204161 | TMEM273 | protein_coding | 0.002823 |
| ENSG00000228340 | MIR646HG | lncRNA | 0.004396 |
| ENSG00000181350 | LRRC75A | protein_coding | 0.004548 |
| ENSG00000198160 | MIER1 | protein_coding | 0.004823 |
| ENSG00000149115 | TNKS1BP1 | protein_coding | 0.005047 |
| ENSG00000179950 | PUF60 | protein_coding | 0.005056 |
| ENSG00000012223 | LTF | protein_coding | 0.005145 |
| ENSG00000053770 | AP5M1 | protein_coding | 0.006177 |
| ENSG00000146426 | TIAM2 | protein_coding | 0.006356 |
| ENSG00000127540 | UQCR11 | protein_coding | 0.006363 |
| ENSG00000105426 | PTPRS | protein_coding | 0.006895 |
| ENSG00000163754 | GYG1 | protein_coding | 0.007322 |
| ENSG00000203485 | INF2 | protein_coding | 0.007612 |
| ENSG00000162894 | FCMR | protein_coding | 0.008494 |
| ENSG00000166341 | DCHS1 | protein_coding | 0.00857 |
| ENSG00000182504 | CEP97 | protein_coding | 0.009035 |
| ENSG00000138639 | ARHGAP24 | protein_coding | 0.009205 |
| ENSG00000114554 | PLXNA1 | protein_coding | 0.010056 |
| ENSG00000116717 | GADD45A | protein_coding | 0.010702 |
| ENSG00000070814 | TCOF1 | protein_coding | 0.012918 |
| ENSG00000181852 | RNF41 | protein_coding | 0.013032 |
| ENSG00000198081 | ZBTB14 | protein_coding | 0.01314 |
| ENSG00000175197 | DDIT3 | protein_coding | 0.013279 |
| ENSG00000136197 | C7orf25 | protein_coding | 0.01368 |
| ENSG00000233038 | PTPRN2-AS1 | lncRNA | 0.013787 |
| ENSG00000102781 | KATNAL1 | protein_coding | 0.014231 |
| ENSG00000156381 | ANKRD9 | protein_coding | 0.014807 |
| ENSG00000033011 | ALG1 | protein_coding | 0.015092 |
| ENSG00000147872 | PLIN2 | protein_coding | 0.016127 |
| ENSG00000183019 | MCEMP1 | protein_coding | 0.016129 |
| ENSG00000171860 | C3AR1 | protein_coding | 0.017092 |
| ENSG00000141456 | PELP1 | protein_coding | 0.017368 |
| ENSG00000110090 | CPT1A | protein_coding | 0.018703 |
| ENSG00000143771 | CNIH4 | protein_coding | 0.018765 |
| ENSG00000134248 | LAMTOR5 | protein_coding | 0.019932 |
| ENSG00000065717 | TLE2 | protein_coding | 0.021082 |
| ENSG00000164818 | DNAAF5 | protein_coding | 0.021283 |
| ENSG00000135452 | TSPAN31 | protein_coding | 0.021379 |
| ENSG00000138069 | RAB1A | protein_coding | 0.021853 |
| ENSG00000101773 | RBBP8 | protein_coding | 0.025976 |
| ENSG00000148450 | MSRB2 | protein_coding | 0.026319 |
| ENSG00000127837 | AAMP | protein_coding | 0.026894 |
| ENSG00000105486 | LIG1 | protein_coding | 0.03201 |
| ENSG00000140545 | MFGE8 | protein_coding | 0.032211 |
| ENSG00000159753 | CARMIL2 | protein_coding | 0.034439 |
| ENSG00000272079 | AC004233.2 | lncRNA | 0.035077 |
| ENSG00000136770 | DNAJC1 | protein_coding | 0.036225 |
| ENSG00000104522 | GFUS | protein_coding | 0.037882 |
| ENSG00000171503 | ETFDH | protein_coding | 0.040553 |
| ENSG00000143387 | CTSK | protein_coding | 0.042935 |
| ENSG00000137460 | FHDC1 | protein_coding | 0.045539 |
| ENSG00000262001 | DLGAP1-AS2 | lncRNA | 0.04916 |
| ENSG00000004660 | CAMKK1 | protein_coding | 0.05188 |
| ENSG00000182310 | SPACA6 | protein_coding | 0.055258 |
| ENSG00000213949 | ITGA1 | protein_coding | 0.057182 |
| ENSG00000242110 | AMACR | protein_coding | 0.057656 |
| ENSG00000121864 | ZNF639 | protein_coding | 0.058361 |
| ENSG00000169372 | CRADD | protein_coding | 0.073386 |
| ENSG00000155090 | KLF10 | protein_coding | 0.092011 |
| ENSG00000143919 | CAMKMT | protein_coding | 0.099486 |
| ENSG00000251136 | AF117829.1 | lncRNA | 0.126656 |
| ENSG00000117133 | RPF1 | protein_coding | 0.186535 |

Data were weights and differential gene expression analysis of 144 gene transcripts included in wb-TRS. Weights were calculated by using a least absolute shrinkage and selection operator logistic-model for non-obese type 2 diabetes and after 10-fold cross-validation in testing dataset. Abbreviation: wb-TRS, whole blood-based transcriptional risk score.

**Table S3.** Receiver operator characteristic curves for prediction of nonobese type 2 diabetes

|  | **Estimate (95% CI)** | **Primary model** | **wb-TRS** | **FBG+TG** | **FBG+TG+ Primary model** | **wb-TRS +**  **Primary model** |
| --- | --- | --- | --- | --- | --- | --- |
| **Training dataset** |  |  |  |  |  |  |
| Primary model | 0.64 (0.60-0.68) |  |  |  |  |  |
| wb-TRS | 0.81 (0.78-0.85) | <.0001 |  |  |  |  |
| FBG+TG | 0.84 (0.80-0.87) | <.0001 | 0.26 |  |  |  |
| FBG+TG+Primary model | 0.85 (0.82-0.88) | <.0001 | 0.08 | 0.27 |  |  |
| wb-TRS+Primary model | 0.82 (0.79-0.85) | <.0001 | 0.41 | 0.33 | 0.10 |  |
| wb-TRS+FBG+TG+Primary model | 0.90 (0.87-0.92) | <.0001 | <.0001 | <.0001 | <.0001 | <.0001 |
| **Testing dataset** |  |  |  |  |  |  |
| Primary model | 0.67 (0.60-0.74) |  |  |  |  |  |
| wb-TRS | 0.78 (0.72-0.84) | 0.01 |  |  |  |  |
| FBG+TG | 0.79 (0.73-0.86) | 0.004 | 0.69 |  |  |  |
| FBG+TG+Primary model | 0.82 (0.77-0.88) | <.0001 | 0.18 | 0.56 |  |  |
| wb-TRS+Primary model | 0.79 (0.72-0.85) | 0.004 | 0.06 | 0.61 | 0.51 |  |
| wb-TRS+FBG+TG+Primary model | 0.86 (0.81-0.91) | <.0001 | 0.0002 | 0.10 | 0.11 | 0.001 |
| **Total dataset** |  |  |  |  |  |  |
| Primary model | 0.65 (0.61-0.68) |  |  |  |  |  |
| wb-TRS | 0.89 (0.87-0.91) | <.0001 |  |  |  |  |
| FBG+TG | 0.82 (0.79-0.95) | <.0001 | 0.001 |  |  |  |
| FBG+TG+Primary model | 0.84 (0.81-0.87) | <.0001 | 0.005 | 0.03 |  |  |
| wb-TRS+Primary model | 0.89 (0.87-0.91) | <.0001 | 0.20 | 0.02 | 0.002 |  |
| wb-TRS+FBG+TG+Primary model | 0.93 (0.91-0.95) | <.0001 | <.0001 | <.0001 | <.0001 | <.0001 |

Estimates (95% CI) were for the area under the receiver operator characteristic curves (AUC). The right numbers were P values for pairwise comparison between the AUC according to the respective models. Primary model comprised age, sex, BMI and family history of diabetes. Abbreviations: CI, confidence interval; wb-TRS, whole blood-based transcriptional risk score; BMI, body mass index; FBG, fasting glucose index; TG, triglycerides.

**Table S4.** Top30 Reactome pathways associated with genes in wb-TRS.

| **Reactome ID** | **Description** | **P value** | **Bonferroni**  **P value** | **Gene** |
| --- | --- | --- | --- | --- |
| R-HSA-381183 | ATF6 (ATF6-alpha) activates chaperone genes | 2.67E-05 | 0.01369 | HSPA5; DDIT3 |
| R-HSA-381033 | ATF6 (ATF6-alpha) activates chaperones | 4.33E-05 | 0.01369 | HSPA5; DDIT3 |
| R-HSA-6804754 | Regulation of TP53 Expression | 9.44E-04 | 0.1983 | TP53 |
| R-HSA-6804114 | TP53 Regulates Transcription of Genes Involved in G2 Cell Cycle Arrest | 0.001728 | 0.26385 | GADD45A; TP53 |
| R-HSA-8941333 | RUNX2 regulates genes involved in differentiation of myeloid cells | 0.002094 | 0.26385 | RUNX2 |
| R-HSA-8941284 | RUNX2 regulates chondrocyte maturation | 0.00283 | 0.29711 | RUNX2 |
| R-HSA-9648895 | Response of EIF2AK1 (HRI) to heme deficiency | 0.004268 | 0.3434 | DDIT3 |
| R-HSA-9031525 | NR1H2 (LXRβ) and NR1H3 (LXRα) regulate gene expression to limit cholesterol uptake | 0.005651 | 0.3434 | MYLIP |
| R-HSA-6791312 | TP53 Regulates Transcription of Cell Cycle Genes | 0.006153 | 0.3434 | GADD45A; TNKS1BP1; TP53 |
| R-HSA-8939246 | RUNX1 regulates transcription of genes involved in differentiation of myeloid cells | 0.006788 | 0.3434 | RUNX2 |
| R-HSA-6799990 | Metal sequestration by antimicrobial proteins | 0.009346 | 0.3434 | S100A8; LTF |
| R-HSA-8941332 | RUNX2 regulates genes involved in cell migration | 0.010761 | 0.3434 | RUNX2 |
| R-HSA-9723907 | Loss of Function of TP53 in Cancer CAPZA1 | 0.010996 | 0.3434 | TP53 |
| R-HSA-9723905 | Loss of function of TP53 in cancer due to loss of tetramerization ability | 0.010996 | 0.3434 | TP53 |
| R-HSA-381042 | PERK regulates gene expression | 0.011686 | 0.3434 | HSPA5; DDIT3 |
| R-HSA-1989781 | PPARA activates gene expression | 0.013749 | 0.3434 | TIAM2; CPT1A; PLIN2 |
| R-HSA-75892 | Platelet Adhesion to exposed collagen | 0.013855 | 0.3434 | ITGA1; GP5 |
| R-HSA-1679131 | Trafficking and processing of endosomal TLR | 0.013855 | 0.3434 | CTSK; LGMN |
| R-HSA-400206 | Regulation of lipid metabolism by PPARalpha | 0.014461 | 0.3434 | TIAM2; CPT1A; PLIN2 |
| R-HSA-2032785 | YAP1- and WWTR1 (TAZ)-stimulated gene expression | 0.017285 | 0.3434 | RUNX2 |

Data were top30 Reactome gene enrichment analyses of 144 gene transcripts included in wb-TRS. Reactome pathway analysis was conducted on reactome website (https://reactome.org/PathwayBrowser/#TOOL=AT). Abbreviation: wb-TRS, whole blood-based transcriptional risk score. **Table S5.** Transcriptome wide Mendelian randomization analysis shows the causal effect of gene transcripts included in wb_TRS on type 2 diabetes.

|  |  | **eQTL →T2D (AGEN)** | | | | **eQTL → T2D (DIAGREM)** | | | |
| --- | --- | --- | --- | --- | --- | --- | --- | --- | --- |
| **Gene_ID** | **Gene_name** | **OR** | **95% CI** | **P** | **SNP_n** | **OR** | **95% CI** | **P** | **SNP_n** |
| ENSG00000179562 | **GCC1-PAX4** | **0.83** | **0.80-0.88** | **5.2×10^-15^** | 2 | 1.01 | 0.95-1.07 | 0.63 | 2 |
| ENSG00000117448 | **AKR1A1** | **0.78** | **0.71-0.86** | **5.0×10^-7^** | 1 | 0.93 | 0.84-1.01 | 0.08 | 1 |
| ENSG00000127837 | **AAMP** | **0.81** | **0.74-0.90** | **3.6×10^-5^** | 1 | 0.87 | 0.79-0.95 | 0.003 | 1 |
| ENSG00000138069 | **RAB1A** | **1.38** | **1.18-1.62** | **8.0×10^-5^** | 1 | 1.47 | 1.21-1.77 | **5.4×10^-5^** | 1 |
| ENSG00000213949 | **ITGA1** | **1.04** | **1.02-1.06** | **0.0002** | 5 | 1.02 | 0.98-1.07 | 0.26 | 8 |
| ENSG00000121864 | **ZNF639** | **1.20** | **1.09-1.35** | **0.0003** | 1 | 1.10 | 0.99-1.22 | 0.06 | 1 |
| ENSG00000143919 | CAMKMT | 0.90 | 0.85-0.97 | 0.004 | 2 | 0.96 | 0.92-0.99 | 0.03 | 4 |
| ENSG00000100211 | CBY1 | 0.91 | 0.87-0.97 | 0.005 | 1 | 0.94 | 0.82-1.08 | 0.37 | 1 |
| ENSG00000130653 | PNPLA7 | 0.96 | 0.95-0.99 | 0.008 | 6 | 1.00 | 0.97-1.02 | 0.74 | 14 |
| ENSG00000069020 | MAST4 | 0.94 | 0.90-0.98 | 0.008 | 4 | 0.99 | 0.95-1.03 | 0.64 | 6 |
| ENSG00000196497 | IPO4 | 1.12 | 1.03-1.23 | 0.01 | 1 | 0.90 | 0.81-0.99 | 0.03 | 1 |
| ENSG00000117133 | RPF1 | 1.58 | 1.10-2.26 | 0.01 | 1 | 1.00 | 0.95-1.05 | 0.99 | 2 |
| ENSG00000102781 | KATNAL1 | 0.93 | 0.87-0.98 | 0.01 | 2 | 1.00 | 0.91-1.10 | 0.96 | 3 |
| ENSG00000124813 | RUNX2 | 0.96 | 0.92-0.99 | 0.02 | 9 | 1.02 | 0.97-1.06 | 0.48 | 11 |
| ENSG00000228340 | MIR646HG | 1.12 | 1.02-1.23 | 0.02 | 2 | 0.97 | 0.87-1.07 | 0.56 | 2 |
| ENSG00000100147 | CCDC134 | 0.94 | 0.88-0.99 | 0.04 | 4 | 1.04 | 0.98-1.11 | 0.17 | 7 |
| ENSG00000091409 | ITGA6 | 0.93 | 0.87-0.99 | 0.04 | 3 | 1.04 | 0.91-1.19 | 0.52 | 2 |
| ENSG00000170345 | FOS | 1.07 | 0.99-1.16 | 0.052 | 4 | 1.09 | 0.95-1.24 | 0.19 | 4 |
| ENSG00000004478 | FKBP4 | 1.05 | 0.99-1.12 | 0.054 | 1 | 1.01 | 0.88-1.14 | 0.91 | 1 |
| ENSG00000110090 | CPT1A | 1.03 | 0.99-1.06 | 0.06 | 3 | 0.97 | 0.94-0.99 | 0.008 | 5 |
| ENSG00000065717 | TLE2 | 1.07 | 0.99-1.14 | 0.07 | 3 | 0.93 | 0.87-0.99 | 0.02 | 3 |
| ENSG00000106991 | ENG | 0.95 | 0.91-1.00 | 0.07 | 2 | 0.97 | 0.94-100 | 0.06 | 4 |
| ENSG00000154358 | OBSCN | 1.08 | 0.99-1.18 | 0.07 | 3 | 1.04 | 0.93-1.14 | 0.47 | 5 |
| ENSG00000183019 | MCEMP1 | 1.04 | 0.99-1.09 | 0.07 | 6 | 1.01 | 0.96-1.05 | 0.7 | 8 |
| ENSG00000135452 | TSPAN31 | 1.02 | 0.99-1.06 | 0.08 | 4 | 1.01 | 0.98-1.04 | 0.41 | 6 |
| ENSG00000132823 | OSER1 | 0.94 | 0.87-1.01 | 0.09 | 8 | 1.06 | 1.03-1.09 | 2.16E-05 | 10 |
| ENSG00000251136 | AF117829.1 | 0.96 | 0.93-1.00 | 0.10 | 5 | 1.01 | 0.97-1.04 | 0.56 | 13 |
| ENSG00000116489 | CAPZA1 | 1.15 | 0.96-1.37 | 0.12 | 1 | 1.14 | 0.98-1.32 | 0.07 | 1 |
| ENSG00000105426 | PTPRS | 1.15 | 0.95-1.39 | 0.14 | 1 | 1.09 | 0.95-1.23 | 0.18 | 2 |
| ENSG00000159753 | CARMIL2 | 0.80 | 0.61-1.07 | 0.14 | 1 | 1.09 | 0.95-1.25 | 0.2 | 1 |
| ENSG00000116815 | CD58 | 1.06 | 0.98-1.14 | 0.14 | 2 | 1.03 | 0.95-1.12 | 0.4 | 3 |
| ENSG00000143771 | CNIH4 | 1.06 | 0.97-1.17 | 0.18 | 4 | 0.98 | 0.96-1.00 | 0.08 | 8 |
| ENSG00000163754 | GYG1 | 1.02 | 0.99-1.07 | 0.18 | 4 | 1.00 | 0.94-1.05 | 0.87 | 4 |
| ENSG00000135049 | AGTPBP1 | 1.02 | 0.99-1.05 | 0.19 | 4 | 1.01 | 0.97-1.04 | 0.6 | 9 |
| ENSG00000106554 | CHCHD3 | 1.08 | 0.96-1.23 | 0.20 | 1 | 1.01 | 0.89-1.14 | 0.88 | 1 |
| ENSG00000179950 | PUF60 | 1.11 | 0.94-1.33 | 0.21 | 1 | 1.06 | 0.86-1.29 | 0.58 | 1 |
| ENSG00000204020 | LIPN | 0.98 | 0.95-1.01 | 0.24 | 10 | 0.99 | 0.96-1.00 | 0.07 | 22 |
| ENSG00000176974 | SHMT1 | 0.98 | 0.96-1.01 | 0.24 | 2 | 0.99 | 0.95-1.02 | 0.41 | 13 |
| ENSG00000114554 | PLXNA1 | 0.95 | 0.89-1.02 | 0.24 | 2 | 1.03 | 0.93-1.14 | 0.54 | 2 |
| ENSG00000214022 | REPIN1 | 1.01 | 0.99-1.05 | 0.25 | 1 | 0.97 | 0.94-1.01 | 0.13 | 1 |
| ENSG00000171503 | ETFDH | 1.01 | 0.99-1.05 | 0.25 | 4 | 1.02 | 0.98-1.05 | 0.25 | 5 |
| ENSG00000225528 | Z82206.1 | 1.03 | 0.98-1.07 | 0.26 | 2 | 1.05 | 1.01-1.09 | 0.005 | 2 |
| ENSG00000157107 | FCHO2 | 1.01 | 0.99-1.05 | 0.27 | 6 | 1.01 | 0.98-1.03 | 0.39 | 15 |
| ENSG00000101773 | RBBP8 | 1.02 | 0.98-1.06 | 0.28 | 6 | 1.02 | 0.98-1.06 | 0.25 | 8 |
| ENSG00000204161 | TMEM273 | 0.97 | 0.92-1.02 | 0.28 | 5 | 1.00 | 0.98-1.01 | 0.83 | 17 |
| ENSG00000143387 | CTSK | 0.96 | 0.93-1.02 | 0.29 | 7 | 0.97 | 0.94-0.99 | 0.01 | 15 |
| ENSG00000149115 | TNKS1BP1 | 1.19 | 0.86-1.63 | 0.29 | 2 | 1.03 | 0.97-1.10 | 0.27 | 3 |
| ENSG00000198160 | MIER1 | 0.98 | 0.95-1.01 | 0.3 | 2 | 1.04 | 1.02-1.07 | 0.0002 | 7 |
| ENSG00000198964 | SGMS1 | 1.91 | 0.91-1.34 | 0.3 | 1 | 0.98 | 0.83-1.16 | 0.8 | 1 |
| ENSG00000182310 | SPACA6 | 1.04 | 0.96-1.13 | 0.31 | 3 | 0.97 | 0.92-1.02 | 0.3 | 4 |
| ENSG00000104522 | GFUS | 1.03 | 0.97-1.09 | 0.33 | 7 | 0.96 | 0.87-1.04 | 0.3 | 10 |
| ENSG00000099337 | KCNK6 | 1.03 | 0.97-1.08 | 0.35 | 5 | 1.03 | 0.99-1.07 | 0.09 | 8 |
| ENSG00000119711 | ALDH6A1 | 0.92 | 0.78-1.09 | 0.36 | 1 | 1.06 | 0.93-1.20 | 0.37 | 1 |
| ENSG00000075826 | SEC31B | 1.01 | 0.98-1.06 | 0.38 | 5 | 1.02 | 0.99-1.04 | 0.12 | 14 |
| ENSG00000164080 | RAD54L2 | 1.08 | 0.90-1.30 | 0.38 | 1 | 0.88 | 0.74-1.04 | 0.14 | 1 |
| ENSG00000140688 | RUSF1 | 0.96 | 0.86-1.05 | 0.38 | 2 | 0.98 | 0.81-1.18 | 0.83 | 2 |
| ENSG00000156381 | ANKRD9 | 0.99 | 0.97-1.01 | 0.39 | 12 | 0.97 | 0.94-0.99 | 0.04 | 17 |
| ENSG00000166527 | CLEC4D | 1.99 | 0.41-9.75 | 0.39 | 2 | 1.48 | 0.77-2.84 | 0.23 | 3 |
| ENSG00000101782 | RIOK3 | 0.95 | 0.87-1.05 | 0.39 | 6 | 0.99 | 0.92-1.06 | 0.73 | 9 |
| ENSG00000166341 | DCHS1 | 0.98 | 0.94-1.02 | 0.4 | 4 | 0.98 | 0.95-1.00 | 0.05 | 7 |
| ENSG00000105486 | LIG1 | 1.01 | 0.98-1.04 | 0.41 | 3 | 1.02 | 0.98-1.05 | 0.35 | 4 |
| ENSG00000143546 | S100A8 | 1.01 | 0.98-1.05 | 0.41 | 2 | 1.01 | 0.96-1.06 | 0.55 | 3 |
| ENSG00000151136 | BTBD11 | 0.99 | 0.96-1.01 | 0.43 | 10 | 0.99 | 0.97-1.01 | 0.3 | 24 |
| ENSG00000175061 | SNHG29 | 0.95 | 0.83-1.08 | 0.43 | 2 | 1.03 | 0.96-1.09 | 0.36 | 3 |
| ENSG00000071051 | NCK2 | 1.02 | 0.97-1.06 | 0.43 | 4 | 1.00 | 0.94-1.05 | 0.88 | 4 |
| ENSG00000148450 | MSRB2 | 1.02 | 0.97-1.07 | 0.44 | 12 | 1.00 | 0.97-1.02 | 0.88 | 21 |
| ENSG00000171860 | C3AR1 | 0.88 | 0.60-1.26 | 0.48 | 3 | 1.06 | 0.90-1.24 | 0.48 | 4 |
| ENSG00000141510 | TP53 | 0.86 | 0.55-1.32 | 0.48 | 1 | 1.02 | 0.91-1.14 | 0.7 | 2 |
| ENSG00000100324 | TAB1 | 1.08 | 0.85-1.38 | 0.49 | 1 | 0.89 | 0.74-1.05 | 0.16 | 1 |
| ENSG00000198663 | C6orf89 | 1.02 | 0.97-1.05 | 0.49 | 1 | 0.98 | 0.92-1.04 | 0.45 | 1 |
| ENSG00000233038 | PTPRN2-AS1 | 0.98 | 0.94-1.02 | 0.50 | 4 | 0.97 | 0.93-1.01 | 0.09 | 5 |
| ENSG00000181852 | RNF41 | 0.87 | 0.58-1.31 | 0.52 | 2 | 0.98 | 0.87-1.08 | 0.57 | 2 |
| ENSG00000063978 | RNF4 | 0.97 | 0.89-1.06 | 0.53 | 3 | 1.03 | 0.96-1.09 | 0.43 | 3 |
| ENSG00000168300 | PCMTD1 | 1.02 | 0.95-1.10 | 0.53 | 2 | 1.02 | 0.97-1.05 | 0.44 | 4 |
| ENSG00000100744 | GSKIP | 0.98 | 0.95-1.02 | 0.53 | 4 | 1.00 | 0.97-1.02 | 0.94 | 4 |
| ENSG00000137460 | FHDC1 | 0.97 | 0.90-1.05 | 0.57 | 2 | 0.99 | 0.90-1.07 | 0.74 | 3 |
| ENSG00000165802 | NSMF | 1.01 | 0.97-1.04 | 0.58 | 6 | 0.96 | 0.92-0.99 | 0.02 | 8 |
| ENSG00000116717 | GADD45A | 0.98 | 0.92-1.05 | 0.59 | 4 | 0.99 | 0.90-1.08 | 0.79 | 5 |
| ENSG00000140545 | MFGE8 | 0.99 | 0.95-1.03 | 0.6 | 6 | 1.00 | 0.98-1.02 | 0.69 | 8 |
| ENSG00000175197 | DDIT3 | 1.02 | 0.95-1.10 | 0.6 | 3 | 1.01 | 0.95-1.07 | 0.79 | 3 |
| ENSG00000162894 | FCMR | 1.02 | 0.95-1.08 | 0.61 | 2 | 1.04 | 0.98-1.10 | 0.13 | 3 |
| ENSG00000170291 | ELP5 | 1.01 | 0.97-1.05 | 0.62 | 2 | 1.06 | 0.99-1.13 | 0.09 | 3 |
| ENSG00000084463 | WBP11 | 1.02 | 0.93-1.11 | 0.62 | 1 | 0.95 | 0.87-1.02 | 0.13 | 1 |
| ENSG00000196565 | HBG2 | 1.01 | 0.97-1.05 | 0.62 | 5 | 1.00 | 0.97-1.02 | 0.69 | 6 |
| ENSG00000117632 | STMN1 | 1.02 | 0.95-1.09 | 0.64 | 2 | 1.06 | 1.00-1.12 | 0.02 | 2 |
| ENSG00000136770 | DNAJC1 | 1.04 | 0.87-1.24 | 0.65 | 2 | 0.99 | 0.90-1.08 | 0.83 | 2 |
| ENSG00000155090 | KLF10 | 0.96 | 0.80-1.15 | 0.67 | 2 | 0.94 | 0.87-1.01 | 0.08 | 2 |
| ENSG00000174791 | RIN1 | 1.00 | 0.98-1.02 | 0.68 | 3 | 1.02 | 0.98-1.06 | 0.26 | 6 |
| ENSG00000146426 | TIAM2 | 1.00 | 0.98-1.03 | 0.68 | 15 | 1.00 | 0.98-1.02 | 0.74 | 18 |
| ENSG00000138639 | ARHGAP24 | 0.99 | 0.97-1.02 | 0.70 | 5 | 1.01 | 0.99-1.04 | 0.21 | 8 |
| ENSG00000004660 | CAMKK1 | 1.03 | 0.88-1.21 | 0.70 | 2 | 0.98 | 0.91-1.05 | 0.55 | 1 |
| ENSG00000152465 | NMT2 | 0.99 | 0.98-1.01 | 0.70 | 6 | 1.00 | 0.98-1.02 | 0.7 | 11 |
| ENSG00000117643 | MAN1C1 | 1.01 | 0.95-1.08 | 0.71 | 6 | 0.96 | 0.81-1.11 | 0.55 | 6 |
| ENSG00000044574 | HSPA5 | 0.97 | 0.82-1.14 | 0.72 | 4 | 0.93 | 0.83-1.04 | 0.18 | 7 |
| ENSG00000125863 | MKKS | 1.01 | 0.97-1.04 | 0.72 | 8 | 1.01 | 0.98-1.04 | 0.32 | 11 |
| ENSG00000178732 | GP5 | 0.98 | 0.90-1.07 | 0.73 | 3 | 0.97 | 0.90-1.05 | 0.48 | 3 |
| ENSG00000198081 | ZBTB14 | 0.99 | 0.93-1.05 | 0.74 | 2 | 1.04 | 0.99-1.08 | 0.07 | 2 |
| ENSG00000203485 | INF2 | 0.99 | 0.96-1.03 | 0.74 | 5 | 1.02 | 0.99-1.05 | 0.08 | 7 |
| ENSG00000173868 | PHOSPHO1 | 1.01 | 0.96-1.05 | 0.75 | 9 | 1.03 | 0.99-1.08 | 0.1 | 11 |
| ENSG00000167074 | TEF | 1.01 | 0.96-1.06 | 0.75 | 6 | 1.02 | 0.97-1.05 | 0.41 | 12 |
| ENSG00000164818 | DNAAF5 | 1.01 | 0.96-1.05 | 0.75 | 2 | 1.00 | 0.96-1.04 | 0.99 | 3 |
| ENSG00000134248 | LAMTOR5 | 1.01 | 0.96-1.06 | 0.76 | 2 | 1.01 | 0.97-1.05 | 0.63 | 3 |
| ENSG00000012223 | LTF | 0.98 | 0.84-1.13 | 0.76 | 6 | 1.00 | 0.94-1.05 | 0.86 | 7 |
| ENSG00000100600 | LGMN | 0.99 | 0.95-1.03 | 0.76 | 4 | 1.00 | 0.95-1.05 | 0.96 | 5 |
| ENSG00000130787 | HIP1R | 0.98 | 0.82-1.15 | 0.78 | 2 | 1.18 | 0.94-1.47 | 0.14 | 2 |
| ENSG00000171456 | ASXL1 | 1.01 | 0.95-1.06 | 0.78 | 1 | 1.02 | 0.97-1.07 | 0.45 | 1 |
| ENSG00000114735 | HEMK1 | 1.01 | 0.96-1.05 | 0.79 | 2 | 1.04 | 0.97-1.11 | 0.23 | 5 |
| ENSG00000147872 | PLIN2 | 1.01 | 0.94-1.08 | 0.79 | 5 | 1.00 | 0.95-1.04 | 0.88 | 12 |
| ENSG00000105447 | GRWD1 | 1.00 | 0.98-1.02 | 0.81 | 1 | 0.99 | 0.92-1.05 | 0.62 | 2 |
| ENSG00000130066 | SAT1 | 1.01 | 0.90-1.14 | 0.82 | 1 | 1.08 | 0.95-1.22 | 0.2 | 1 |
| ENSG00000143797 | MBOAT2 | 0.99 | 0.88-1.10 | 0.82 | 1 | 1.01 | 0.93-1.09 | 0.84 | 2 |
| ENSG00000007944 | MYLIP | 1.01 | 0.89-1.15 | 0.84 | 6 | 1.04 | 0.89-1.20 | 0.63 | 8 |
| ENSG00000178904 | DPY19L3 | 1.00 | 0.96-1.04 | 0.85 | 2 | 1.05 | 1.00-1.10 | 0.02 | 2 |
| ENSG00000106665 | CLIP2 | 1.00 | 0.96-1.04 | 0.86 | 8 | 1.04 | 0.99-1.08 | 0.1 | 8 |
| ENSG00000112335 | SNX3 | 0.98 | 0.82-1.18 | 0.86 | 1 | 0.99 | 0.87-1.12 | 0.86 | 2 |
| ENSG00000053770 | AP5M1 | 1.01 | 0.87-1.17 | 0.88 | 1 | 1.10 | 0.95-1.26 | 0.2 | 1 |
| ENSG00000262001 | DLGAP1-AS2 | 1.01 | 0.92-1.09 | 0.88 | 2 | 1.04 | 0.97-1.11 | 0.27 | 2 |
| ENSG00000172725 | CORO1B | 0.99 | 0.93-1.06 | 0.89 | 4 | 1.02 | 0.98-1.06 | 0.33 | 5 |
| ENSG00000160013 | PTGIR | 0.99 | 0.91-1.08 | 0.89 | 5 | 0.99 | 0.92-1.05 | 0.67 | 6 |
| ENSG00000182504 | CEP97 | 1.00 | 0.94-1.07 | 0.89 | 2 | 0.99 | 0.90-1.09 | 0.88 | 2 |
| ENSG00000148803 | FUOM | 1.01 | 0.87-1.15 | 0.93 | 3 | 0.97 | 0.92-1.01 | 0.15 | 6 |
| ENSG00000242110 | AMACR | 1.00 | 0.96-1.03 | 0.94 | 3 | 0.98 | 0.95-1.01 | 0.2 | 7 |
| ENSG00000136197 | C7orf25 | 1.00 | 0.98-1.02 | 0.96 | 8 | 0.98 | 0.96-1.00 | 0.06 | 12 |
| ENSG00000102908 | NFAT5 | 1.00 | 0.75-1.33 | 0.97 | 2 | 1.17 | 0.73-1.86 | 0.51 | 2 |
| ENSG00000181350 | LRRC75A | 1.00 | 0.92-1.07 | 0.97 | 3 | 0.98 | 0.87-1.09 | 0.66 | 5 |

Data were odds ratio (OR) and 95% confidence interval (CI). Transcriptome wide Mendelian randomization was used to evaluate causal effects of genes transcripts included in wb_TRS on type 2 diabetes. Abbreviation: eQTL, expression quantitative trait locus; AGEN, the Asian Genetic Epidemiology Network; DIAGREM, the DIAbetes Genetics Replication And Meta-analysis.

**Fig S1.** Flowchart of study participants.

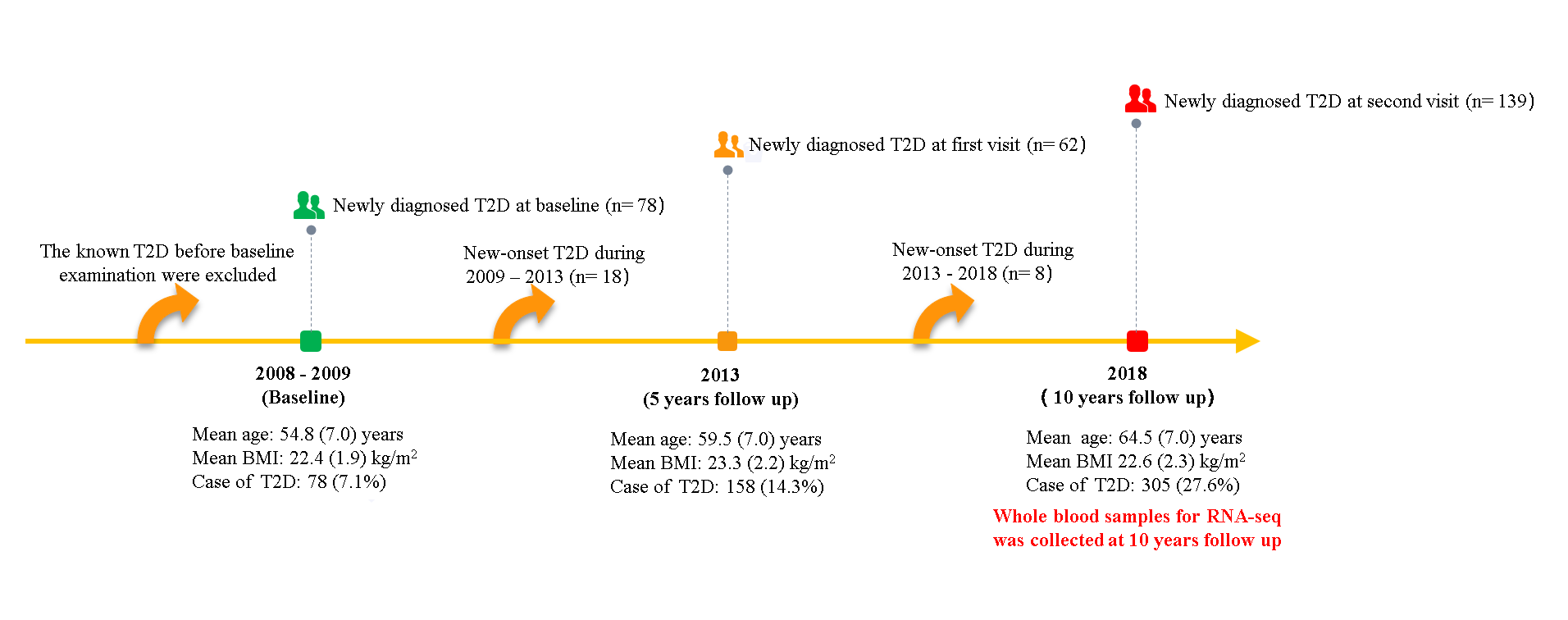

**Fig S2.** Cross-validation results for least absolute shrinkage and selection operator (LASSO) logistics-model for nonobese type 2 diabetes in training dataset.

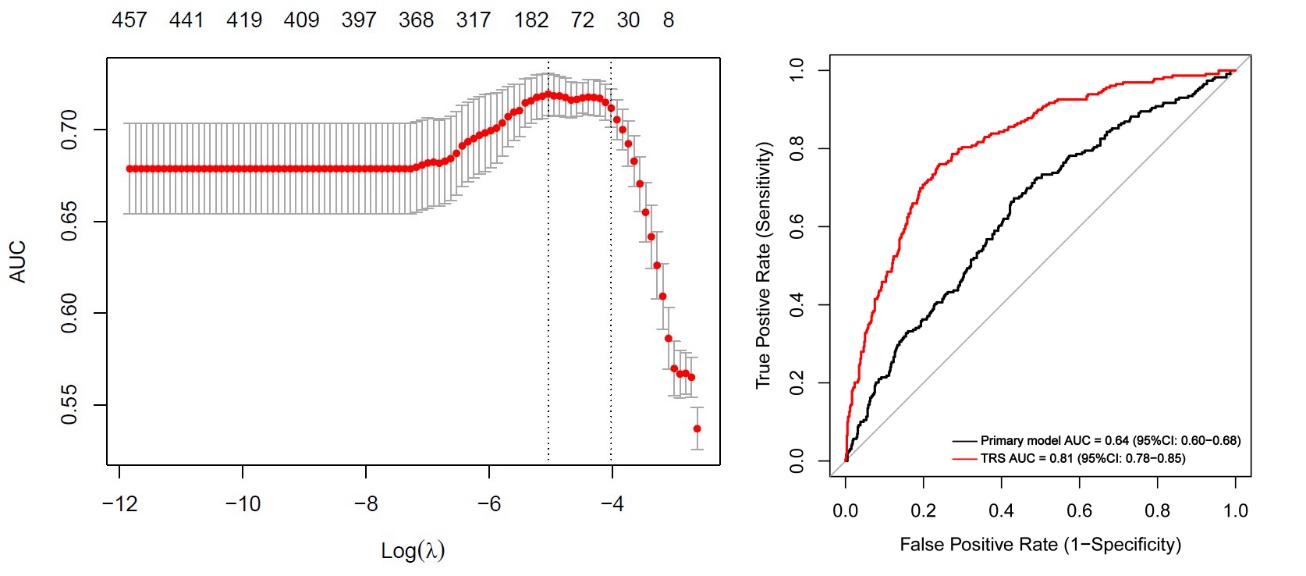

**Fig S3.** Violin plot of wb-TRS and gene transcripts most negatively (FUOM) and positively (RPF1) associated with nonobese type 2 diabetes.

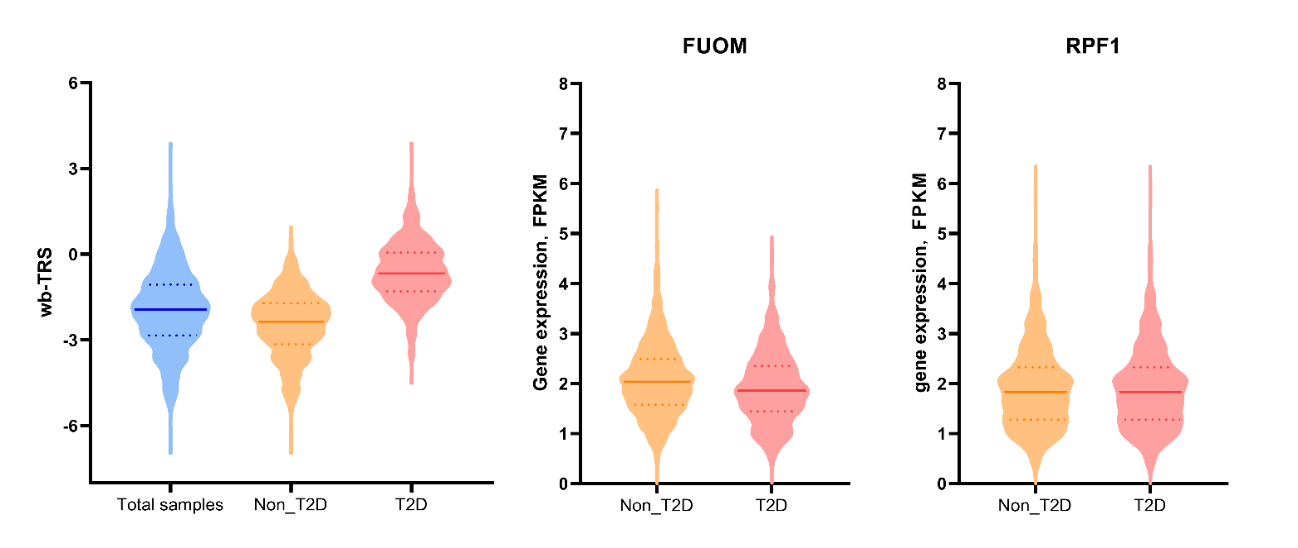

**Fig S4.** Stratified analysis.

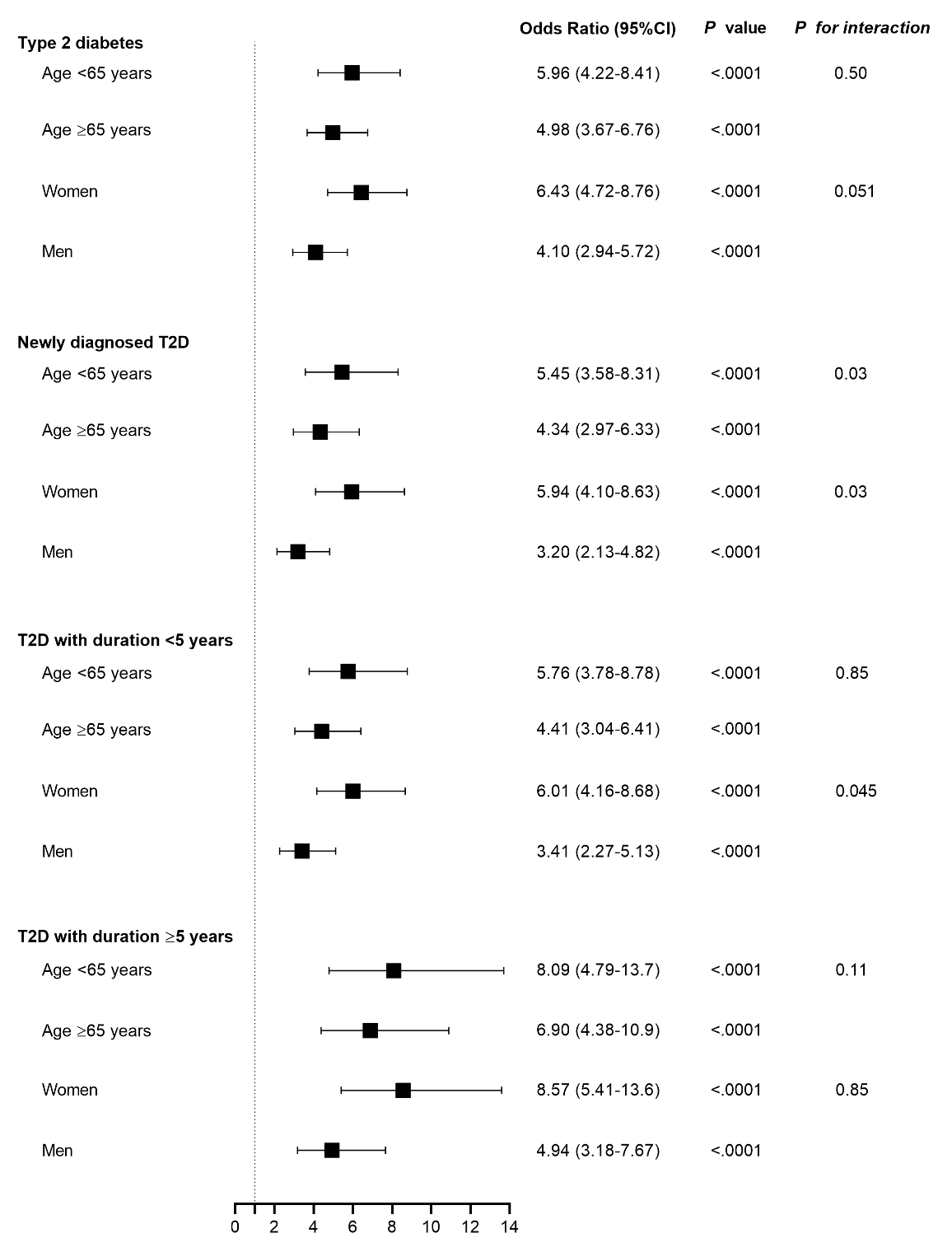
